# Positive selection underlies repeated knockout of ORF8 in SARS-CoV-2 evolution

**DOI:** 10.1101/2023.09.21.23295927

**Authors:** Cassia Wagner, Kathryn E. Kistler, Garrett A. Perchetti, Noah Baker, Lauren A. Frisbie, Laura Marcela Torres, Frank Aragona, Cory Yun, Marlin Figgins, Alexander L. Greninger, Alex Cox, Hanna N. Oltean, Pavitra Roychoudhury, Trevor Bedford

**Affiliations:** Department of Genome Sciences, University of Washington, Seattle, WA, USA; Vaccine and Infectious Disease Division, Fred Hutchinson Cancer Center, Seattle, WA, USA; Howard Hughes Medical Institute, Seattle, WA, USA; Department of Laboratory Medicine and Pathology, University of Washington, Seattle, Washington, USA; Washington State Department of Health, Shoreline, Washington, USA; Department of Applied Mathematics, University of Washington, Seattle, Washington, USA

## Abstract

Knockout of the ORF8 protein has repeatedly spread through the global viral population during SARS-CoV-2 evolution. Here we use both regional and global pathogen sequencing to explore the selection pressures underlying its loss. In Washington State, we identified transmission clusters with ORF8 knockout throughout SARS-CoV-2 evolution, not just on novel, high fitness viral backbones. Indeed, ORF8 is truncated more frequently and knockouts circulate for longer than for any other gene. Using a global phylogeny, we find evidence of positive selection to explain this phenomenon: nonsense mutations resulting in shortened protein products occur more frequently and are associated with faster clade growth rates than synonymous mutations in ORF8. Loss of ORF8 is also associated with reduced clinical severity, highlighting the diverse clinical impacts of SARS-CoV-2 evolution.

## Introduction

Selection pressure on SARS-CoV-2 has shaped the population of circulating virus since its emergence in humans. The virus has undergone repeated selective sweeps of variant of concern viruses, such as Delta and Omicron, and more recently by lineages within-Omicron, including BA.2 and XBB, in which increased fitness derives from mutations contributing to both intrinsic transmissibility and immune escape^1–11^. Adaptive mutations are overrepresented in spike, the viral entry protein and primary target of protective adaptive immunity, and mutations here alter tropism, improve transmission, and evade host immunity^12–17^. The number of mutations in S1, the spike subunit containing the receptor binding domain, correlate with viral growth rate^18^.

Adaptive evolution has not been limited to spike, however. Specific missense mutations in open reading frames (ORFs) for non-structural (ORF1a and ORF1b), other structural (nucleocapsid, N) and accessory (ORF3a) proteins are also associated with increased viral fitness^19,20^. ORF8 has repeatedly been knocked out during SARS-CoV-2 evolution, though the evolutionary pressures acting on loss of ORF8 are not known. Multiple large deletions of ORF8 and occasionally neighboring ORF7a and ORF7b have been identified around the world, including in Singapore in 2020, where it was associated with reduced clinical severity^21–25^. Additionally, premature stops in ORF8 causing early truncation of the 121-amino acid protein have been reported, including in mink and pangolin animal species, the Alpha variant of concern (Q27*) and lineage XBB.1 descendants (G8*)^3,11,26–28^. As of September 2023, the vast majority (∼90%) of currently circulating SARS-CoV-2 has ORF8 knocked out^29^. This pattern mirrors SARS-CoV’s loss of ORF8 after introduction into humans^30^.

ORF8 is a viral accessory protein that aids in immune evasion^31^. As a secreted protein, it drives an early antibody response^32,33^, potentially acting as a decoy for protective adaptive immunity. Many functions have been attributed to ORF8, including downregulating major histocompatibility complex class I (MHC I) which decreases antibody dependent cellular cytoxicity activity^33–36^, inhibiting Type I IFN production^37–40^, suppressing IFN-γ induced antiviral gene expression^41^, and disrupting host epigenetic regulation by acting as histone H3 mimic ^42^. In its unconventional, unglycosylated state, ORF8 may contribute to cytokine storms by activating the IL-17 pathway^43–45^.

Given these varied potential functions of ORF8, its repeated knockout is perplexing. One hypothesis is that ORF8 knockout is deleterious to SARS-CoV-2 fitness but rose to fixation frequency by hitchhiking along with fitness enhancing mutations in Alpha and again in XBB.1 descendants. Another hypothesis is that ORF8 knockout has no impact on viral fitness, and the gene is undergoing neutral evolution. Here, again, fixation could be explained by hitchhiking. A final hypothesis is that ORF8 knockout improves viral fitness, and positive selection for knockout has contributed to its global spread.

To explore these hypotheses, we use SARS-CoV-2 sequences from Washington State (WA) from February 2020-March 2023 to determine prevalence of ORF8 knockout across time, contrasting this with the knockout of other SARS-CoV-2 genes. Here, we can observe knockouts occurring on a variety of fitness backgrounds, not just the fit viral backbones which swept globally. Next, we use a large, global phylogeny of SARS-CoV-2 to compare expected counts and clade growth rates of nonsense mutations, which truncate the ORF8 protein, to synonymous mutations in ORF8. Finally, we assess linked hospitalization and death data to determine the clinical impact of ORF8 knockout.

## Results

We quantified how often ORF8 was knocked out during SARS-CoV-2 evolution in WA from the beginning of the COVID-19 pandemic through March 2023. Our dataset included knockouts under a wide potential array of selection pressures, including knockouts which primarily spread locally and knockouts in the Alpha and XBB.1 descendant viruses which spread globally. As the first U.S. state to detect community transmission of SARS-CoV-2, WA has robustly sequenced COVID-19 cases throughout the pandemic aided by a sentinel surveillance sequencing system for geographic coverage^46–48^. From April 2021 through March 2023, 17.25% of all COVID-19 infections in WA were sequenced, with the lowest sequencing coverage in December 2021 (3% of cases) and the highest in February 2022 (28% of cases). This high sequencing coverage makes WA an ideal location to understand prevalence of ORF8 knockout across time^49^.

We considered samples to contain a potential knockout in ORF8 if they contained a large deletion (>30 bp) or a premature stop codon resulting in a 10 codons shorter protein coding sequence. This cutoff, though arbitrary, prevents mislabelling common, short deletions as knockouts while avoiding to preferentially maximize or minimize knockouts in any one gene (Fig S1). Samples with a mutation known to cause amplicon dropout in ORF8 were excluded from potential knockouts (see methods). We identified 14,929 samples with a potential knockout of ORF8, representing 11.7% of high coverage (>=95%) SARS-CoV-2 sequences collected in WA through March 2023 (Fig 1A). For ORF8, the number of knockouts was robust to cutoff length: with a cutoff of 95 codons missing, 9.9% of sequences would still have an ORF8 knockout (Fig S1).

**Fig 1.**
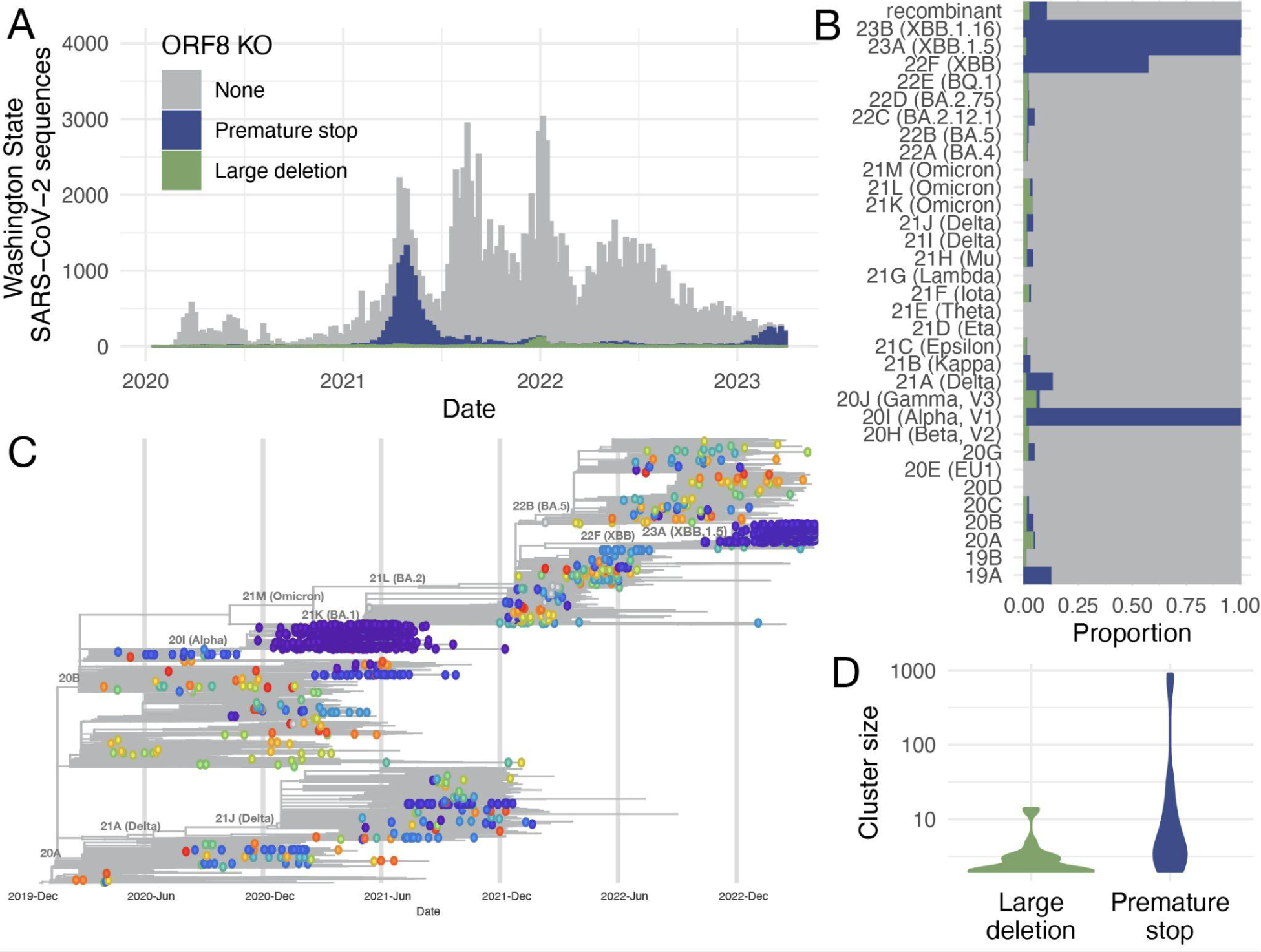
ORF8 is repeatedly knocked out during SARS-CoV-2 evolution in Washington State. (A) Distribution of the number of SARS-CoV-2 sequences collected in WA by collection date. Histogram is colored by the type of potential ORF8 knockout (none=gray, premature stop = blue, large deletion = green). (B) Proportion of sequences with a potential ORF8 knockout by Nextstrain Clade. (C) Time-resolved phylogenetic tree of 16,268 SARS-CoV-2 sequences enriched for sequences in WA (9,854) evenly sampled across time through March 2023. Tips with a potential ORF8 knockout are shown as circles colored by a unique cluster. There are 355 unique clusters, so colors are reused, but adjacent tips of the same color belong to the same cluster. All other tips are plotted as gray lines. (D) Violin plots of cluster size for ORF8 knockouts due to large deletions (green) or premature stops (green).

While the majority of ORF8 knockouts were found in variants descending from clade 20I (Alpha), clade 22F (lineage XBB), clade 23A (lineage XBB.1.5), and clade 23B (lineage XBB.1.16), ORF8 knockout also occurred in an average of 3% of all other clades (Fig 1B). Most knockouts were due to premature stop codons, either from nonsense or frameshift mutations, with only 10.2% of knockouts being large deletions. This suggests that most knockouts are real and not artifactual errors in sequencing as point mutations and small gaps can be confidently inferred with short read sequencing and reference-based genome assembly.

We constructed a phylogenetic tree enriched for sequences sampled in WA spread evenly across time to determine if potential knockouts clustered together phylogenetically. We identified parsimony clusters of ORF8 knockouts across the tree using unidirectional clustering for large deletions and bidirectional clustering for premature stops (see methods) (Fig 1C). We identified 355 unique clusters: 250 large deletion clusters and 105 premature stop clusters. Most clusters were singletons, with only 53 clusters containing at least two samples. Premature stop clusters were larger with a mean cluster size of 17.2 compared to 1.2 for large deletions (*p*=2.3e-04, Wilcoxon Rank Sum test) (Fig 1D). This difference in cluster size could reflect different fitnesses associated with different types of gene knockout. For example, multiple non-singleton, large deletion clusters had deletions over 300bp, which resulted in knockout of both ORF8 and ORF7b. However, among non-singleton clusters, deletion size was positively correlated with cluster size (Pearson’s *r* = 0.47, *p*=0.012) (Fig S2A). More likely the difference in cluster size by knockout type occurs because many potential large deletions represent a sequencing error, rather than a large deletion, and fail to cluster with other potential large deletions.

To determine whether potential large deletions were true deletions versus amplicon dropouts or sequencing errors, we screened a subset using PCR and Sanger sequencing. Of 9998 University of Washington samples available at the time of screening, 120 were found to have sequences with contiguous strings of Ns (>266 bp) from ORF7a through ORF8. Of these, 89 samples had sufficient volume and quality for PCR and Sanger sequencing, and 23/89 (25.8%) were confirmed to have large deletions (≥344 bp) (Table S1).

For knockouts with premature stop codons, we did not find a correlation between truncated protein length and cluster size (Pearson’s *r* = –0.14, *p* = 0.49) (Fig S2B). However, 77% of non-singleton knockout clusters due to an early stop mutation were predicted to have a truncated protein of 26 codons or smaller (Fig S2D). This skewed distribution alone suggests a non-random process underlying premature stop codons.

Next, we compared the number of potential ORF8 knockouts to potential knockout of other genes in SARS-CoV-2 in our WA dataset. With 13,410 premature stop codons, ORF8 had 14x more stop codons than any other gene (Fig 2A). The largest genes, ORF1a, ORF1b, and Spike, contained the most large deletions, with >24,000 in each compared to 1,517 large deletions in ORF8 (Fig S3A). Given the necessity of these genes to viral replication, this finding suggests that many potential large deletions represent missing bases due to poor sequence coverage rather than true deletions. Analyzing the constituent proteins of the ORF1a & ORF1b genes did not show any evidence of a deletion hotspot relative to other SARS-CoV-2 proteins (Fig S3C). When normalized by gene length, ORF7b had the highest number of large deletions (Fig S3B). However, we observe that non-singleton knockout clusters in ORF8 (mean 34.3) are larger on average than non-singleton knockout clusters for all other genes (mean 3.1) (*p*=1.4e-05, Wilcoxon Rank Sum test) (Fig 2B). This result is driven by premature stop clusters (Fig S3C), as we detect no significant difference in large deletion cluster size among genes with the largest deletion cluster sizes: spike (mean 3.3), ORF8 (mean 3.4) and M (mean 4.1) (ANOVA, *p* = 0.09) (Fig S3D). These results are consistent with ORF8 being knocked out more frequently than any other gene in SARS-CoV-2, and the difficulty of identifying large deletions from assembled sequences.

**Fig 2.**
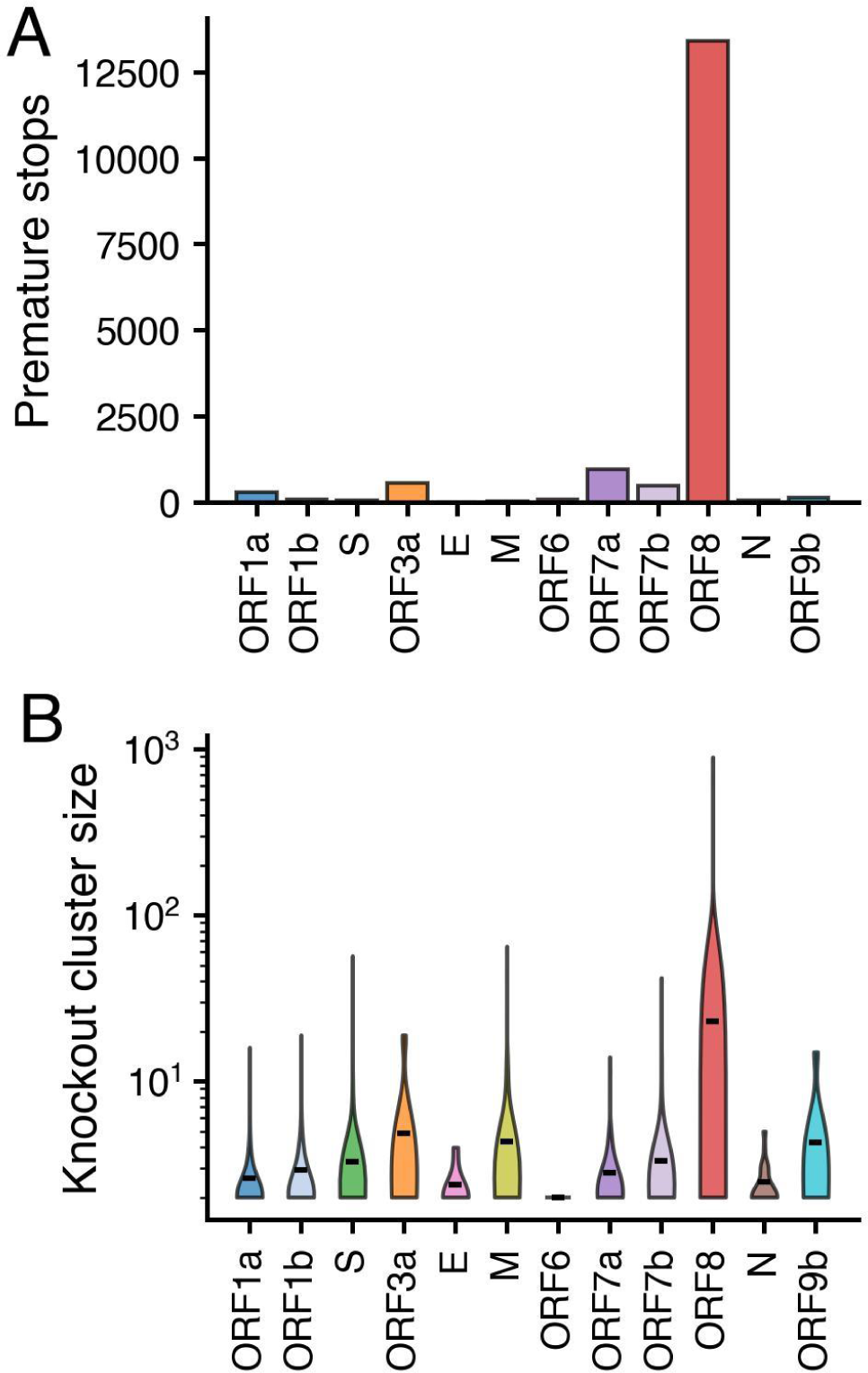
ORF8 has more premature stops and larger knockout clusters than any other gene. (A) Number of premature stops by gene in WA SARS-Cov-2 sequences through March 2023. (B) Size of parsimony clusters with a gene knockout due to large deletion or premature stop for all SARS-CoV-2 genes. Clusters were reconstructed from the maximum likelihood phylogenetic tree enriched for WA sequences with even temporal sampling.

In WA, we observed that ORF8 is truncated more commonly than any other gene, and clusters containing an ORF8 knockout are larger than clusters with a knockout in any other gene. This result suggests either weakened selection pressure on maintaining ORF8 function relative to other genes or positive selection for ORF8 knockout. To better test these hypotheses, we applied one of the most widely used measures of selection pressure, *dN*/*dS*, which compares the ratio of mutation divergence over expectation for both nonsynonymous, or protein modifying, mutations and synonymous mutations, which do not alter the protein’s amino acid sequence. Classically, *dN*/*dS*>1 is consistent with positive selection as nonsynonymous mutations occur more frequently than synonymous mutations, *dN*/*dS*<1 is consistent with negative selection, and *dN*/*dS* ∼ 1 is consistent with neutral evolution. Here, we modified the classic calculation of *dN*/*dS* to separately estimate values for missense mutations, which change the amino acid sequence, and nonsense mutations, which introduce a stop codon and result in early truncation of the protein. To mitigate geographic bias, we estimated *dN*/*dS* values for each SARS-CoV-2 gene using the publicly available UShER phylogeny, which contains ∼7 million SARS-CoV-2 sequences sampled from around the globe (Fig 3A, S4A, S5) ^50,51^. For clarity, we focus the main text on ORF8, comparing results with spike, which has undergone substantial adaptive evolution, and the more conserved ORF1a, a polyprotein cleaved into 10 nonstructural proteins. A single nonsense mutation in ORF1a could knockout multiple constituent proteins, so we chose to examine the gene as a whole.

**Fig 3.**
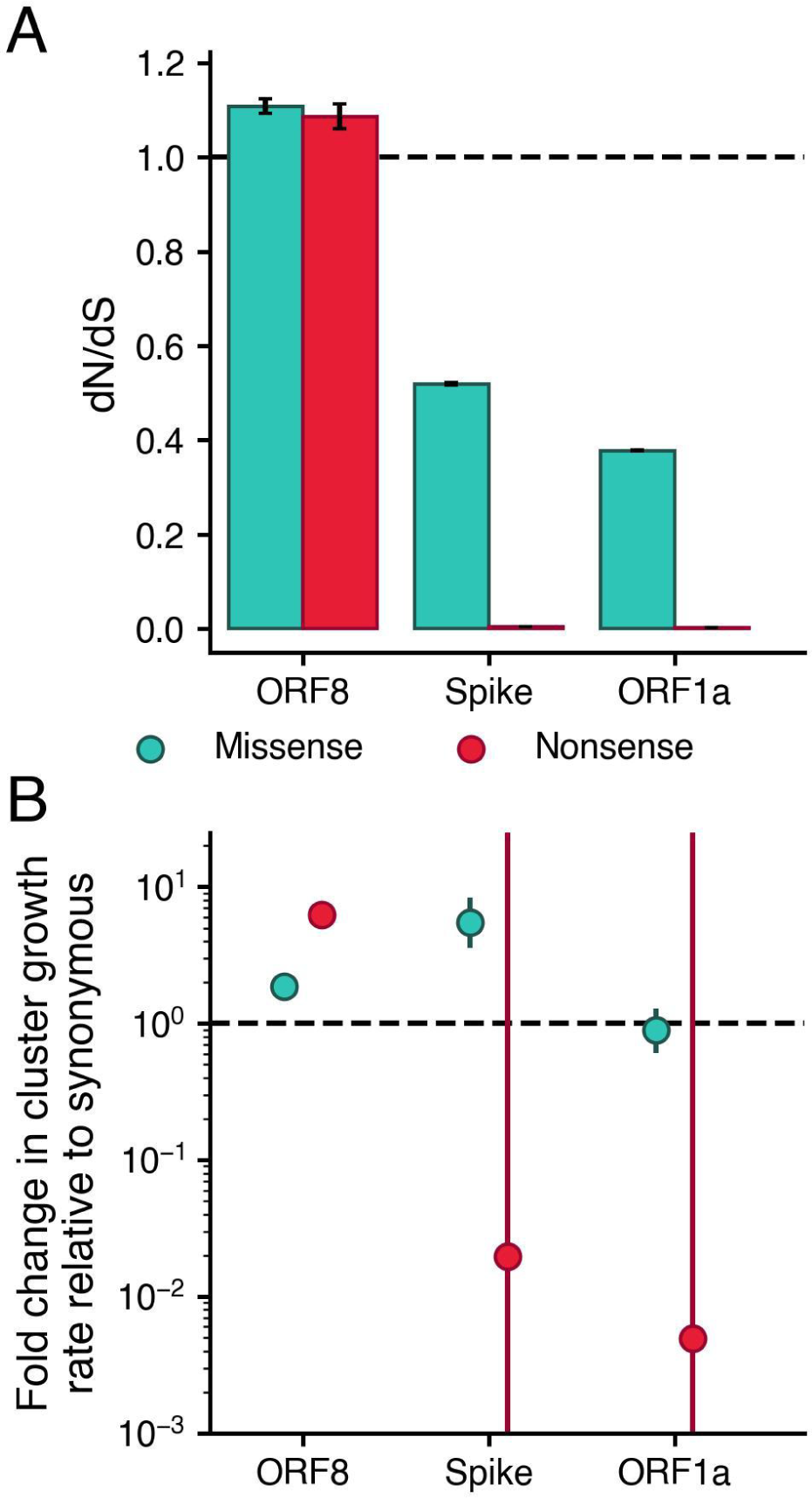
Nonsense mutations in ORF8 result in faster clade growth rates and are more frequently observed than synonymous mutations. From the global, UShER SARS-CoV-2 phylogeny, we estimated (A) dN/dS and (B) the fold change in mutation cluster growth rate relative to synonymous for missense (teal) and nonsense (red) mutations. Error bars show 95% confidence intervals. For dN/dS, confidence intervals were calculated by 10,000 bootstrap iterations across nodes in the tree.

In spike and ORF1a, we identify strong selection against nonsense mutations, with *dN*/*dS* values <0.01. This result is consistent with both genes being necessary for viral replication. In spike, selection against missense mutation is relaxed relative to ORF1a (with *dN/dS* values of 0.52 and 0.38 respectively), which is consistent with well-documented adaptive evolution in spike^14,18^. In comparison in ORF8, we observe much higher *dN*/*dS* values for both missense and nonsense mutations (1.09 and 1.11 respectively). Classically, *dN*/*dS* values of this magnitude are consistent with positive selection; however, caution is warranted when interpreting within-population *dN/dS* in terms of selective coefficients^52,53^. Additionally, absolute *dN/dS* values are sensitive to substitution matrix used while relative relationships of estimates between genes remain similar regardless of substitution matrix (Fig S4). Comparing across genes, we can conclude that negative selection on ORF8 is strongly weakened relative to spike and ORF1a. Our results further suggest positive selection for ORF8 knockout: even with an alternative substitution matrix, *dN/dS* estimates for ORF8 remained >1 (Fig S4).

To more clearly test for positive selection, we compared success of ORF8 clades with a nonsense mutation to clades with either a synonymous or missense mutation in ORF8 (Fig S6). We found that clades with a nonsense mutation in ORF8 are larger (mean = 77.6, std = 6024.2) and circulate for longer (mean = 11.5 days, std = 35.8) than clades with a synonymous mutation in ORF8 (mean size = 7.0, std = 423.8, mean days = 9.5, std = 28.9). Clades with a missense mutation in ORF8 are also larger on average (mean = 18.5, std = 2482.0) and circulate for longer (mean = 10.3, std = 32.1). For comparison, nonsense mutations in ORF1a and spike are much smaller and circulate for far shorter periods than clades with synonymous mutations.

To statistically quantify these observed differences, we modeled the rate of cluster growth by mutation type as a negative binomial regression of the number of descendants after the mutation was first observed, with an offset for time since observation (Fig 3B). We found that clusters with nonsense mutations in ORF8 grow 6.3x (95% CI: 5.97-6.52) faster than clusters with synonymous mutations in ORF8. While this approach does not attempt to disentangle the effects of other fitness-impacting mutations which occur downstream of a nonsense mutation, the synonymous cluster growth rate provides a null expectation for comparison. Assuming an absence of epistatic interactions between ORF8 nonsense mutation and other fitness enhancing mutations in SARS-CoV-2, this result suggests that the observed ORF8 knockouts boost viral fitness. This effect is robust to excluding nonsense mutations found in Alpha and XBB descendants, which occurred on highly fit backbones: nonsense mutations still grew 2.5x (95% CI: 2.30-2.76) faster than clusters with synonymous mutations (Table S2). Missense mutations in ORF8 grew 1.8x faster (95% CI: 1.77 to 1.96) than clusters with synonymous mutations in ORF8. This relative fitness benefit could result from either (1) missense mutations disrupting ORF8 function like nonsense mutations, or (2) missense mutations improving fitness by enhancing some aspect of ORF8 function.

In comparison, clusters with a nonsense mutation in spike and ORF1a grew at 0.02x and 0.005x the rate of clusters with a synonymous mutation. However, these differences were not statistically significant (*p*=0.49 and *p*=0.47 respectively), likely due to the very few number of nonsense mutation clusters observed resulting in wide CIs (Fig 3B). In spike and ORF1a, 0.035% and 0.017% of mutations were nonsense respectively. Comparing cluster growth rates conditions on mutations occurring and being sampled, so rates will be less sensitive for detecting clusters with mutations under strong negative selection.

The difference in growth rate with a nonsense mutation in ORF8 is similar to that of increase in growth rate for a missense mutation in spike: 5.5x (95% CI: 3.57-8.38). Since many missense mutations in Spike are associated with fitness gains, this finding also suggests positive selection for ORF8 knockout. We did not observe a significant difference in growth rate for missense mutations in ORF1a (0.88, 95% CI: 0.607-1.29). However, if mutations are split out into mutations with positive fitness previously inferred by a hierarchical logistic regression model^19^ versus other missense mutations, clusters with fitness-associated missense mutations grew 4.3x faster (95% CI: 2.08-10.11) than synonymous mutation clades while cluster with other missense mutations grew 0.43x slower (95% CI: 0.29-0.64x) relative to synonymous (Fig S7). This is consistent with predominantly negative selection on ORF1a, but occasional mutations under positive selection. We observed a similar split of cluster growth rates for ORF8 and spike when classifying mutations by type and previously inferred positive fitness. These results suggest that our cluster growth analysis is in agreement with previous work (Fig S7)^19^. Calculated growth rates by mutation type for all other genes are available in the supplement (Fig S8).

Previous analysis found a large deletion that knocked out ORF8 and truncated ORF7b to be associated with reduced clinical severity^22^. Here, we decided to extend this analysis to the clinical impact of any ORF8 knockout, by a large deletion or premature stop codon, by linking SARS-CoV-2 sequences with clinical outcomes recorded in the Washington Disease Reporting System. Given the reduced clinical severity and loss of vaccine efficacy associated with Omicron variants, we restricted our analysis to pre-Omicron lineages. Table S3 outlines the general characteristics of our study population stratified by infections with and without an ORF8 knockout. While 8.3% (n=1906/22,928) of individuals infected by SARS-CoV-2 with intact ORF8 were hospitalized, only 6.7% (n=383/5,746) of individuals infected by virus with ORF8 knocked out were hospitalized (*p* = 4e-05, *χ*^2^ test) (Fig 4A). Similarly, 1.8% (n=910/49,912) of individuals infected by virus with intact ORF8 died due to SARS-CoV-2 compared to 1.3% (n=129/10,089) of individuals infected by virus with an ORF8 knockout (*p* = 1.6e-04, *χ*^2^ test) (Fig 4A). However, ORF8 knockouts have not been distributed evenly across time in Washington State (Fig 1A), and clinical severity of SARS-CoV-2 has varied temporally with changing age circulation patterns, the rollout of vaccines, accumulation of natural immunity in the population, medications, and viral evolution.

**Fig 4.**
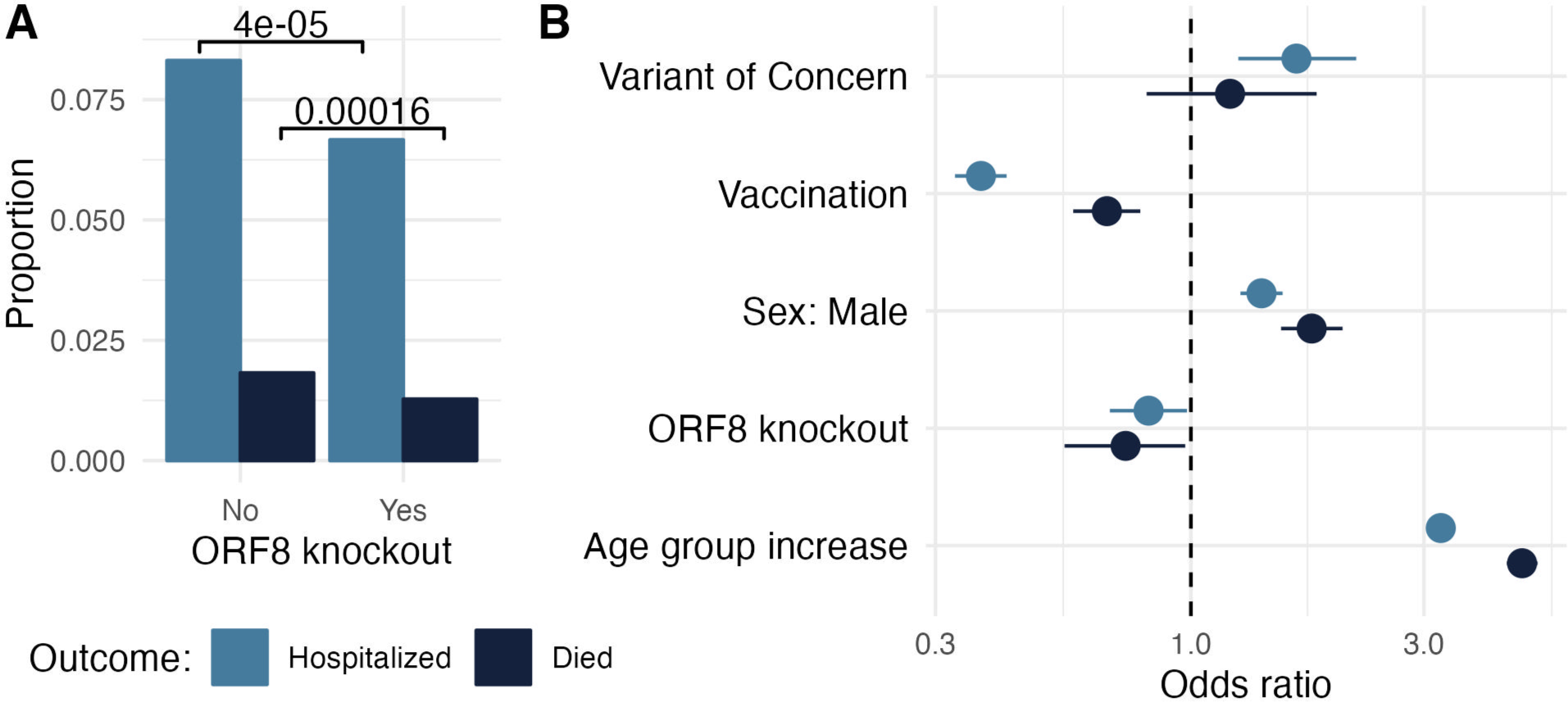
ORF8 knockout is associated with reduced clinical severity of COVID-19. (A) Proportion of individuals with severe COVID-19 outcomes stratified by virus infection with and without ORF8 knockout. P-values for ɑ=0.05 from χ^2^ test are shown. (B) Odds ratios from a generalized linear model of clinical outcomes for variant of concern (Alpha, Beta, Gamma or Delta), vaccination status, assigned male sex at birth, ORF8 knockout, and increasing age. Bars show 95% confidence intervals. Severe COVID-19 outcomes are hospitalization (light blue) and death (dark blue). Analysis was limited to pre-Omicron lineages due to reduced clinical severity and loss of vaccine efficacy associated with Omicron variants.

In a general linear model adjusting for week of collection, variant of concern, vaccination status, sex at birth, and age group, we found a 0.82 (95% CI: 0.68-0.98) odds ratio of hospitalization in infections containing an ORF8 knockout compared to infections without the knockout (Fig 4B). The odds of death when infected by virus with an ORF8 knockout was also reduced (Odds ratio: 0.73, 95% CI: 0.55-0.97). In both regressions, vaccination was associated with reduced clinical severity while male sex and increase in age group were associated with worse clinical outcomes. When compared to other SARS-CoV-2 lineages, variants of concern – Alpha, Gamma, Delta, or Beta lineages – were independently associated with increased odds of hospitalization but not with odds of death.

Given the difficulty of accurately calling large deletions in ORF8, we tested the robustness of our effects by size of cluster required to define a knockout. To calculate cluster size, we built three additional maximum likelihood phylogenies enriched for ORF8 knockouts in WA one for Delta, one for Alpha, and one for other non-Omicron lineages. These breakdowns were chosen such that all ORF8 knockouts sequenced in WA could be placed in an appropriate phylogenetic context of at least 75% background sequences. We then reconstructed parsimony clusters for ORF8 knockout (see above and Methods). We found that both effect size for ORF8 knockout and model Akaike information criterion (AIC) minimally changed with various cluster sizes required to define a knockout (S6). This demonstrates that the clinical effect is robust to inaccurate identification of ORF8 knockout due to large deletion.

## Discussion

The SARS-CoV-2 pandemic has been characterized by a high rate of evolution as fitness enhancing mutations, primarily in spike, have repeatedly swept globally. Here, we explored the selection pressures underlying a surprising and repeated sweeping mutation pattern: ORF8 knockout.

Examining ORF8 knockout across time in a Washington State, we found that while knockout spread widely in Alpha and XBB.1 descendant lineages, it also occurred at a low frequency on many other viral backbones due to both large deletions and premature stop codons (Fig 1). This finding is consistent with other reports of large deletions encompassing ORF8 circulating in other parts of the globe^21,23–27^. While knockout is observed in other genes in Washington State^54^, we find that ORF8 has more premature stops than any other gene and knockout clusters with ORF8 grow larger than knockout clusters for any other gene (Fig 2). At a global level, we estimate a higher than expected number of nonsense and missense mutations in ORF8 (Fig 3). Nonsense mutations in ORF8 show the highest nonsynonymous over synonymous divergence for any gene in SARS-CoV-2.

Together these results recommend rejecting our first hypothesis: that ORF8 knockout is deleterious to SARS-CoV-2 fitness and fixation was driven by hitchhiking on the back of other fitness enhancing mutations. The *dN/dS*=1.1 estimates suggest that ORF8 knockout is due to positive selection, though estimates in a single evolving population should be interpreted with caution^52,53^. We next modeled the rate of mutation cluster growth rates and found that clusters with nonsense mutations grow roughly 6x faster than synonymous mutations in ORF8. Excluding stop mutation clusters present in XBB and Alpha, we still find that nonsense mutations in ORF8 grow 2.5x faster than synonymous. These values are comparable to the improvement in cluster growth rates by missense mutations in spike over synonymous and further suggest that ORF8 knockout boosts viral fitness.

This conclusion is broadly consistent with other estimates of the fitness effects of SARS-CoV-2 mutations. In an updated run of the fitness model presented in Obermeyer et al, numerous stop mutations in ORF8 are estimated to boost fitness^19^. Bloom and Neher only find evidence of relaxed purifying selection on ORF8 and other SARS-CoV-2 accessory proteins. However, the difference between their fitness effects and our *dN/dS* estimates for ORF8 are consistent with the limited correlation between fitness effects and *dN/dS* estimates they observed. As count-based methods, both approaches are underpowered to identify positive selection since they only explore how often mutation occurs, not how large clades get once mutation occurs^20^. We also found evidence of relaxed purifying selection in SARS-CoV-2 accessory proteins as we estimated high nonsense and missense dN/dS values for ORF7a and ORF7b, in addition to ORF8 (Fig S4). However, unlike ORF8, we did not find evidence for a growth rate advantage for ORF7a or ORF7b knockouts (Fig S7). This clarifies previous reports of ORF7a and ORF7b deletions^21,24,25,54^, and our observation that ORF8 knockout clusters grew larger than for any gene. It also suggests that ORF7a and ORF7b could be deleted in future SARS-CoV-2 evolution. However, ORF8 may be deleted more quickly, due to the fitness benefit associated with ORF8 knockout.

Consistent with previous analysis^22^, we found evidence of reduced clinical severity with ORF8 knockout. This observation might help explain why Alpha had reduced clinical severity compared to the Beta, Gamma, and Delta variants of concern^47^. It also highlights the importance of studying the clinical impact of SARS-CoV-2 evolution; genetic changes in the virus can have different effects on clinical severity.

Our results imply that SARS-CoV-2 genomic surveillance should include detection of ORF8 knockout going forward. Currently, much of circulating SARS-CoV-2 has ORF8 knocked out; however, rescue of ORF8 protein expression could increase the clinical severity of COVID-19 infections. Conversely, ORF8 knockout arising on viral backgrounds with intact ORF8 expression would suggest a transmission advantage. Knockout due to point mutation or frameshifts can be readily detected from assembled viral genomes. To detect large deletions, assembled genomes can be screened for long stretches of N’s, which will result in numerous false positives, or raw reads and sequence alignment maps can be screened for ORF8 deletions earlier in genome assembly pipelines.

A dispensable ORF8 and increased viral replication speed due to a shortened genome cannot explain positive fitness effects associated with premature stop codons. Recent work by Kim et al identifies an intriguing biological mechanism underlying positive selection for ORF8 knockout and the timeline for knockout to sweep^55^. Their study finds that ORF8 covalently interacts with spike at the endoplasmic reticulum, reducing onward transport of spike to the cell membrane and incorporation into virus particles. Presence of ORF8 is associated with less spike in pseudovirions. Less spike in virions and on the cell surface might improve viral fitness within the individual by providing another mechanism for SARS-CoV-2 to avoid the host immune response. However, when ORF8 is knocked out, more spike in virions might improve viral fitness at a transmission level by making it easier to establish infection. This tug of war between within-host fitness and between-host fitness could underlie the length of time it has taken for ORF8 knockout to fix relative to other fitness boosting mutations in SARS-CoV-2. Since SARS-CoV also lost ORF8^30^, and it would be interesting to explore if ORF8 plays a similar role in SARS-CoV.

## Materials and Methods

### Dataset & code availability

On April 24, 2023, we downloaded all 149,547 SARS-CoV-2 sequences from GISAID collected in WA through March 31, 2023^56^. This dataset was used to identify knockouts in ORF8 in WA and to build WA focused phylogenies. We also used an additional 32,363 SARS-CoV-2 sequences sampled elsewhere in the United States and around the globe sampled prior to March 21, 2023 and downloaded from GISAID on July 27, 2023 as contextual sequences in phylogenies. All GISAID metadata and sequences used in analyses are available at gisaid.org/EPI_SET_230921by.

For selection analyses, to mitigate geographic bias, we downloaded the publicly available, mutation-annotated, SARS-CoV-2 UShER tree ^50,51^ on May 1, 2023 from: http://hgdownload.soe.ucsc.edu/goldenPath/wuhCor1/UShER_SARS-CoV-2/. For our analyses, we trimmed this tree to remove sequences without associated collection dates using matUtils 0.6.2 (https://usher-wiki.readthedocs.io/en/latest/matUtils.html#).

All code used in this analysis is available at: https://github.com/blab/ncov-orf8. Code was written in both Python 3.10.9 and R 4.1.2.

### Calling gene knockouts

Sequences were called as having a potential knockout in a gene if either 30 consecutive nucleotide bases in the coding sequence for that gene were gap characters or N’s, or if the predicted protein coding sequence was more than 10 codons shorter than the reference protein, due to a premature stop codon from a nonsense or frameshift mutation. With short-read sequencing and reference-based genome assembly as are commonly used in SARS-CoV-2 sequencing pipelines^57^, large deletions will show up as long stretches of N’s; however, long stretches of N’s could also represent poor sequence quality or amplicon dropout. To limit bias from poor sequencing quality, samples had to have a genome coverage of at least 95%, or no more than 1,495 missing bases. In ORF8, we excluded calling large deletions between bp 27809-27854 in samples with a C27807T mutation as this mutation was associated with amplicon dropout in the ARTIC v4 sequencing primers^58^. We considered alternative cutoff lengths for knockouts, balancing between wrongly calling short, likely functional deletions as gene knockouts and preferentially maximizing or minimizing the number of knockouts in any one gene (Fig S1).

### Sanger sequencing & PCR validation of large deletions

We performed screening by PCR and Sanger sequencing on a subset of samples to determine whether long strings of ambiguous bases (Ns) in ORF7 and ORF8 were the result of deletions rather than amplicon dropout. All 9,998 samples sequenced by the University of Washington as of May 2021 were screened, and 120 were found to have sequences with contiguous strings of Ns (>266 bp) from ORF7a through ORF8. Of these 120, 89 were determined to have sufficient volume and sample quality for PCR and Sanger Sequencing. Total nucleic acid extraction was done on the MagNA Pure 96 instrument (Roche) with 200µL sample input and 50µL elution volume. Amplification was performed with SuperScript-III One-Step RT-PCR kit (Invitrogen, Waltham, MA, USA) and primers designed flanking the deletion region, beginning in ORF7a (forward: GGCACTGATAACACTCGCTAC) through the beginning of the N-gene (reverse: GAGGGTCCACCAAACGTAATG). Thermocycling conditions were as follows: 55°C for 30 min, 94°C for 2 min, and 35 cycles of 94°C for 30 sec, 58°C for 30 sec, and 68°C for 1 min. A final extension step was included at 72°C for 5 min. Reactions were cleaned with SPRI Ampure beads (Beckman Coulter, Brea, CA, USA) at a 0:0.65 volumetric ratio and eluted to 40µL. Flash gel electrophoresis (Lonza, Basel, Switzerland) was performed to confirm successful PCR and for preliminary deletion calling. Samples were diluted and sent for Sanger sequencing on ABI’s Prism 3730xl DNA analyzer (Genewiz, Seattle, WA, USA) with the designed primers. Consensus sequences were aligned against NC_045512.2 to confirm the presence of the deletions.

### Phylogenetic reconstructions

To determine if potential gene knockouts might be part of the same transmission cluster, we built a maximum likelihood phylogeny of SARS-CoV-2 enriched for WA sequences. We used the Nextstrain pipeline^59^ to align sequences to Wuhan-Hu-1/2019 (genbank accession MN908947.3) using nextalign^60^, to reconstruct a maximum-likelihood phylogeny using IQ-TREE^61^, to estimate molecular clock branch lengths using TreeTime^62^, and to infer nucleotide and amino acid substitutions across the phylogeny. The input to the pipeline was a focal ∼10,000 sequences collected in WA and an additional ∼10,000 contextual sequences – 5,000 sequences from the rest of the United States and 5,000 sequences from other countries. For each geographic region, all sequences were sampled evenly across time from the beginning of the SARS-CoV-2 pandemic through March 2023. We used the default settings for the Nextstrain SARS-CoV-2 pipeline (https://github.com/nextstrain/ncov/tree/master) to filter this dataset, except we increased genome coverage >= 95% to minimize large deletions that represent poor sequence coverage. The pipeline additionally excludes samples with incomplete dates, samples with >20 deviation from the molecular clock rate, samples with >5 private reversions, and samples with more than 6 private mutations in a 100-nucleotide window. The final phylogeny contained 16,268 sequences. This phylogeny is available to view at: https://nextstrain.org/groups/blab/ncov/WA/20k.

Also using the Nextstrain pipeline, we built three additional clade-specific phylogenies enriched for sequences with potential large deletions in ORF8 sampled in WA. We built one for Alpha, one for Delta, and one with all other, pre-Omicron SARS-CoV-2 lineages. These numbers were chosen such that every sequence collected in WA with a potential knockout of ORF8 was contained in a phylogeny. The input to the pipeline was all potential ORF8 KO’s in that SARS-CoV-2 clade, no more than 5,000 sequences. For context, we included 5,000 additional WA sequences without ORF8 KO’s, 5,000 sequences from the rest of the United States, and 5,000 sequences from around the globe. Contextual samples were evenly temporally sampled from each geographic region. The pipeline filtered samples as above, and the final trees respectively included 18,350, 14,908, 12,050 sequences. These phylogenies are available to view at: https://nextstrain.org/groups/blab/ncov/WA/Alpha, https://nextstrain.org/groups/blab/ncov/WA/Delta, and https://nextstrain.org/groups/blab/ncov/WA/other.

### Knockout clustering

Knockout clusters were called using clustering methods appropriate for each knockout type. Large deleted segments can only be recovered via recombination, and for the purpose of this analysis we considered this an unlikely event. Therefore, clusters of large deletions were reconstructed using the Camin-Sokal parsimony algorithm^63^, which is a unidirectional parsimony clustering algorithm. We considered sequences to be part of the same deletion cluster if all their common ancestor nodes and all descendant nodes shared a deleted region of at least 30 nucleotides. Premature stop codons introduced by nonsense or frameshift mutations can be removed by back mutation. Thus, knockout clusters due to premature stops were called using the Fitch parsimony algorithm, which allows for back mutation^57,64^. Samples were considered as part of the same knockout cluster if all their common ancestor nodes contained the same premature stop codon.

### Identifying mutation clusters

To compare cluster size and circulation time by mutation type for SARS-CoV-2 genes, we classified point mutations on each node in the SARS-CoV-2 UShER tree as synonymous, missense, or nonsense. Nodes containing multiple mutations in the same gene were excluded from the analysis. Cluster size represented the total number of tips descended from that node and days of circulation was calculated from the latest to the earliest date at which a descendant tip was sampled. By this definition, clusters may contain nested clusters with mutations in that gene that could contribute to their success. However, we chose this definition as using non-nested clusters with a single mutation type per gene truncates signals of positive selection because cluster size is maxed out as a function of the molecular clock rate and length of the gene.

### Calculating dN/dS

We calculated the expected number of synonymous, missense, and nonsense sites for each gene by multiplying each base in the coding sequence by the substitution rates for that base previously inferred for SARS-CoV-2. We then summed together expected mutations by mutation type for each gene. We considered two sources for inferred substitution rates: (1) substitution rates calculated from the 4-fold degenerate sites within SARS-CoV-2 using the global UShER phylogeny^20^, and (2) a maximum likelihood substitution matrix inferred early in the pandemic from 36 SARS-CoV-2 genomes^65^. In the main text, we present results from the first source (Fig 3A), but include results for all genes for both substitution matrices in the Supplement (S4).

We calculated the observed number of synonymous, missense and nonsense mutations in the UShER phylogeny by classifying the reconstructed mutations at each node by their gene and mutation type. We generated the divergence for each mutation type for each gene by dividing the number of observed mutations by the expected number of sites. For missense and nonsense mutations, *dN/dS* were calculated by dividing the divergence for those respective mutation types by the synonymous divergence. While the signal strength observed with an UShER tree as opposed to a smaller global phylogeny is weakened since the number of segregating polymorphisms in the population is greatly increased^52^, our analysis focused on comparing the relative differences in *dN/dS* by mutation type across genes rather than the absolute magnitude of *dN/dS* values.

### Modeling mutation cluster growth rate

Using negative binomial regression, we can model number of additional descendants, given we observed a mutation as:

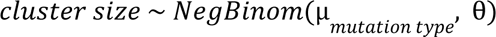

Where µ_*mutation type*_ is the expected number of descendants of a given mutation and θ is the over-dispersion relative to Poisson distribution. Since each cluster can grow for a different period of time, we can model number of additional descendants per unit time since the mutation was first observed by:

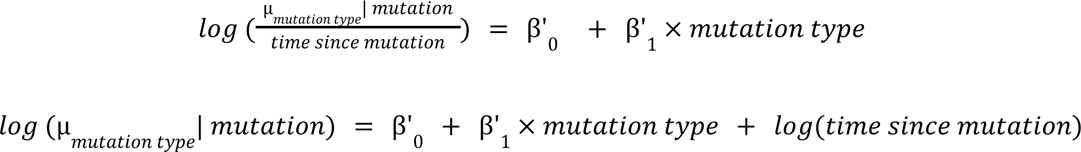

The estimated parameters β’_1_ then correspond to the log fold increase in the cluster size growth rate for a given mutation.

Likelihood ratio test for negative binomial regression compared to Poisson regression indicated that the negative binomial model was more likely due to overdispersion of cluster growth rate (*p*=0). The negative binomial model was fit in R using the MASS package (https://cran.r-project.org/web/packages/MASS/index.html), and the Poisson model was fit in R using the GLM package (https://cran.r-project.org/web/packages/glm2/glm2.pdf), specifying family = Poisson. We fit negative binomial models separately for each gene with mutation types split out by synonymous, missense, and nonsense. For Spike, ORF1a, and ORF8, we fit additional models with mutations split out by type and fitness advantage previously inferred by a hierarchical logistic regression model ^19^.

### Clinical analysis

Under Washington State IRB Exempt Determination 2020-102, age, sex, hospitalization, death, and vaccination history was provided by the Washington Department of Health from the Washington Disease Reporting System for individuals with linked sequenced SARS-CoV-2 samples from June 1, 2020 through July 31, 2022. We limited the clinical analysis to pre-Omicron lineages since Omicron was associated with reduced clinical severity and loss of vaccine efficacy.

We used a *χ*^2^ test to compare the number of individuals who were hospitalized or died due to SARS-CoV-2 infection by presence of an ORF8 knockout in their sequenced sample.

To estimate the impact of ORF8 knockout on clinical outcomes of hospitalization and death, we used a multivariate logistic regression:

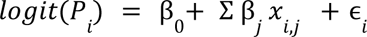

Where *P* is the probability of hospitalization or death, *β* is the coefficient of the predictor variable, *x* is the predictor variable, and *ϵ* is the residual error. Predictor variables were: ORF8 knockout (binary variable), sex at birth (binary variable), age group (discrete variable), vaccinated (binary variable, variant of concern (binary variable), and week of collection (categorical variable). Only sex at birth: Male or Female were included in the model as there were to few Other samples to estimate a coefficient. Age groups were 0-4yo, 5-17yo, 18-44yo, 45-65yo, 65-79yo, and 80+yo. Variants of concern were Alpha, Beta, Delta, and Gamma lineages as designated by the World Health Organization^66^. Indivduals were considered to be vaccinated if two weeks passed since any COVID-19 vaccination. The model was fit in R using the GLM package (https://cran.r-project.org/web/packages/glm2/glm2.pdf).The package Glm was used to conduct the logistic regression. To identify the power to determine a significant effect of ORF8 knockout on death, we used the pwr package in R (https://www.rdocumentation.org/packages/pwr/versions/1.3-0/topics/pwr-package). Specifically, we used power test for two proportions given the number of deaths in our dataset by presence or absence of an ORF8 knockout and the effect size calculated by the GLM.

Given the challenges of classifying ORF8 knockouts due to large deletions, we explored robustness of our model fit and effect size by the criteria required to classify an ORF8 knockout. We introduced the additional criteria that an ORF8 knockout had to cluster with some threshold number of other ORF8 knockout samples in order to be considered a true knockout. We then computed Akaike Information Criterion and the odds ratio of ORF8 knockout for outcomes of hospitalization and death using thresholds from 0-50.

## Data Availability

All GISAID metadata and sequences used in analyses are available at gisaid.org/EPI_SET_230921by. The SARS-CoV-2 UShER phylogeny used in this study was downloaded on May 1, 2023 from http://hgdownload.soe.ucsc.edu/goldenPath/wuhCor1/UShER_SARS-CoV-2/. Clinical data from the Washington Disease Reporting System is not shared due to patient confidentiality. All code used in this analysis is available at: https://github.com/blab/ncov-orf8.

https://www.epicov.org/epi3/epi_set/EPI_SET_230921by?main=true

https://github.com/blab/ncov-orf8

## Acknowledgements

We would like to thank Allison Thibodeau, Topias Lemetyinen, Allison Warren, Cameron Ashton, Emily Nebergall, Peter Gibson, and Sarah Menz for their work throughout the pandemic linking SARS-CoV-2 sequences in Washington State to the Washington Disease Reporting System.

We also gratefully acknowledge all data contributors, i.e., the Authors and their Originating laboratories responsible for obtaining the specimens, and their Submitting laboratories for generating the genetic sequence and metadata and sharing via the GISAID Initiative, on which this research is based.

## Author contributions

CW designed and performed all analyses. KK provided key support for selection analyses. GAP, NB identified and confirmed initial ORF8 deletions. LAB and LMT were critical to clinical data collection and curation. LAB and HNO provided key support for clinical analyses. FA and CY linked clinical metadata to sequences. MF provided key support for cluster growth rate analysis. AC, HNO oversaw clinical data collection and curation. ALG, HNO, PR, and TB oversaw the study. CW and TB wrote the manuscript. All other authors edited the manuscript.

## Supplemental

**S1.**
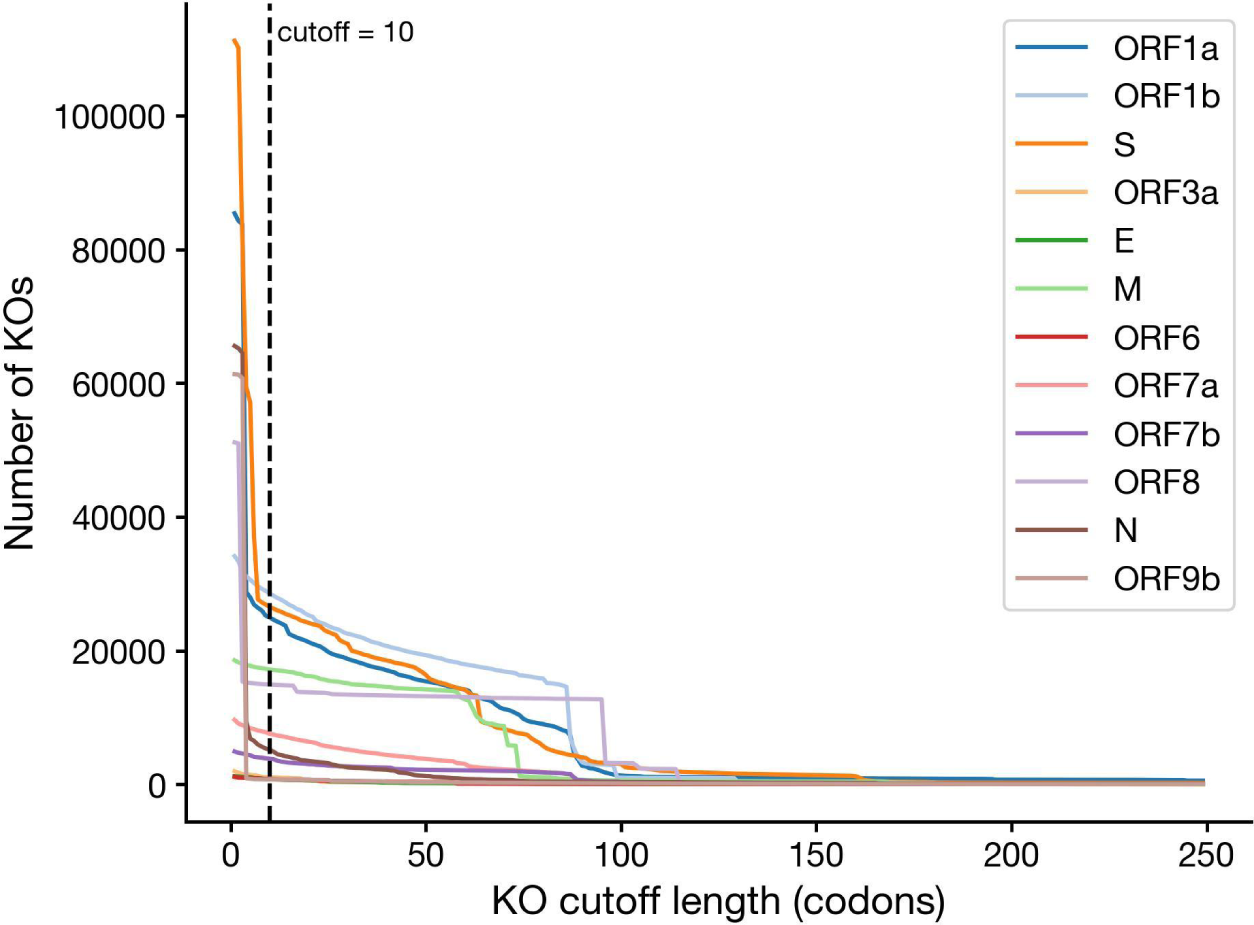
Number of knockouts in WA SARS-CoV-2 sequences by knockout cutoff length. Here, we show the impact of alternative cutoffs to define a gene knockout. Knockout cutoff length refers to the total number of codons that would be missing given a large deletion or premature stop. The dashed vertical line shows the cutoff used in our analysis: 10 codons missing or 30 continuous N or gap characters.

**S2.**
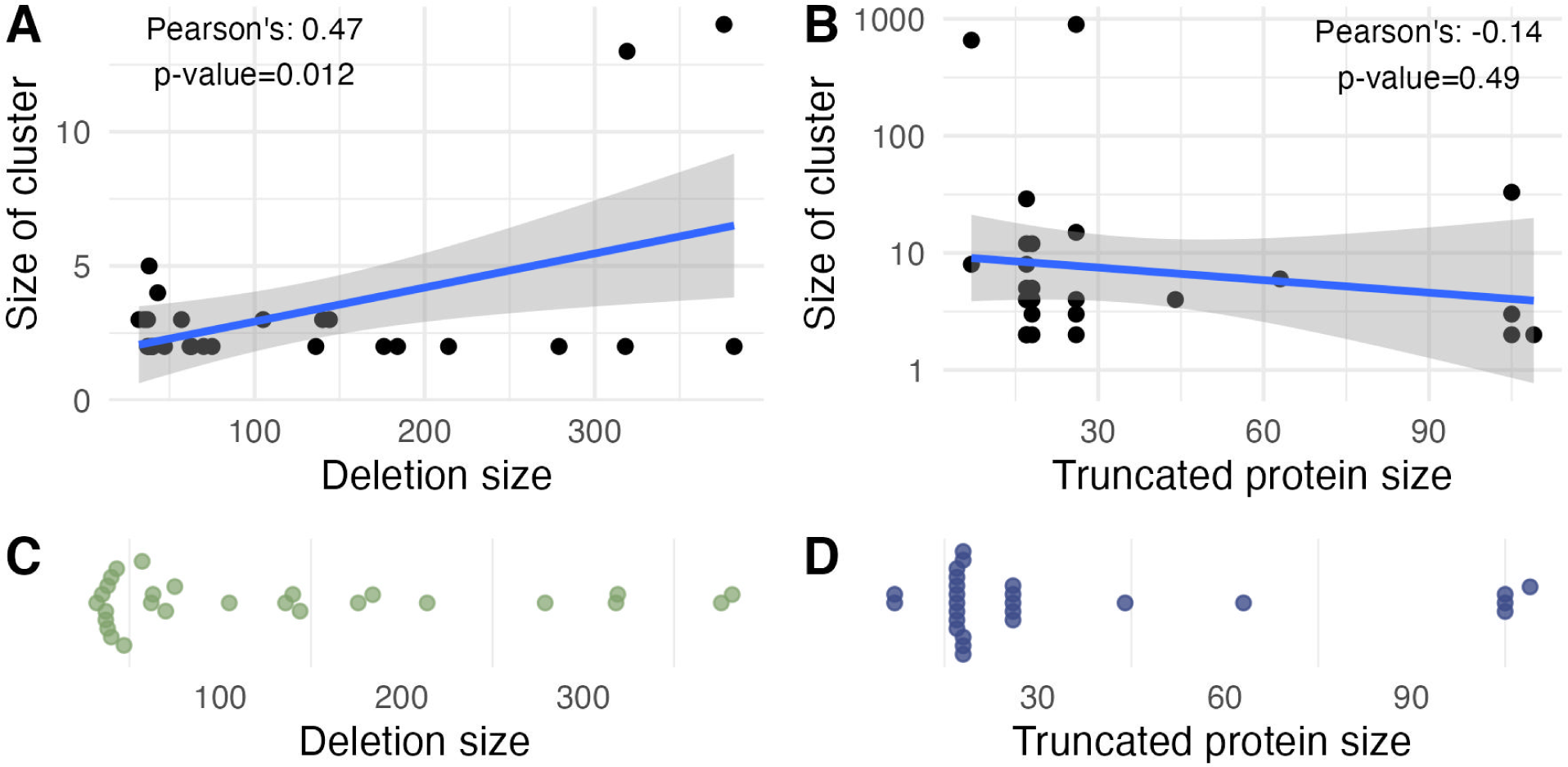
Distribution of deletion size, truncated protein size, and cluster size for phylogenetic clusters with an ORF8 knockout. Here, we show the correlation between (A) deletion size (bp) and cluster size and (B) truncated protein size (codons) and cluster size (B) for phylogenetic clusters with a knockout in an ORF8. (C) Distribution of deletion size in bp for large deletion ORF8 knockout clusters. (D) Distribution of truncated protein sizes in codons for premature stop ORF8 knockout clusters. Clusters were reconstructed from the Washington state SARS-CoV-2 phylogeny shown in Fig 1C.

**S3.**
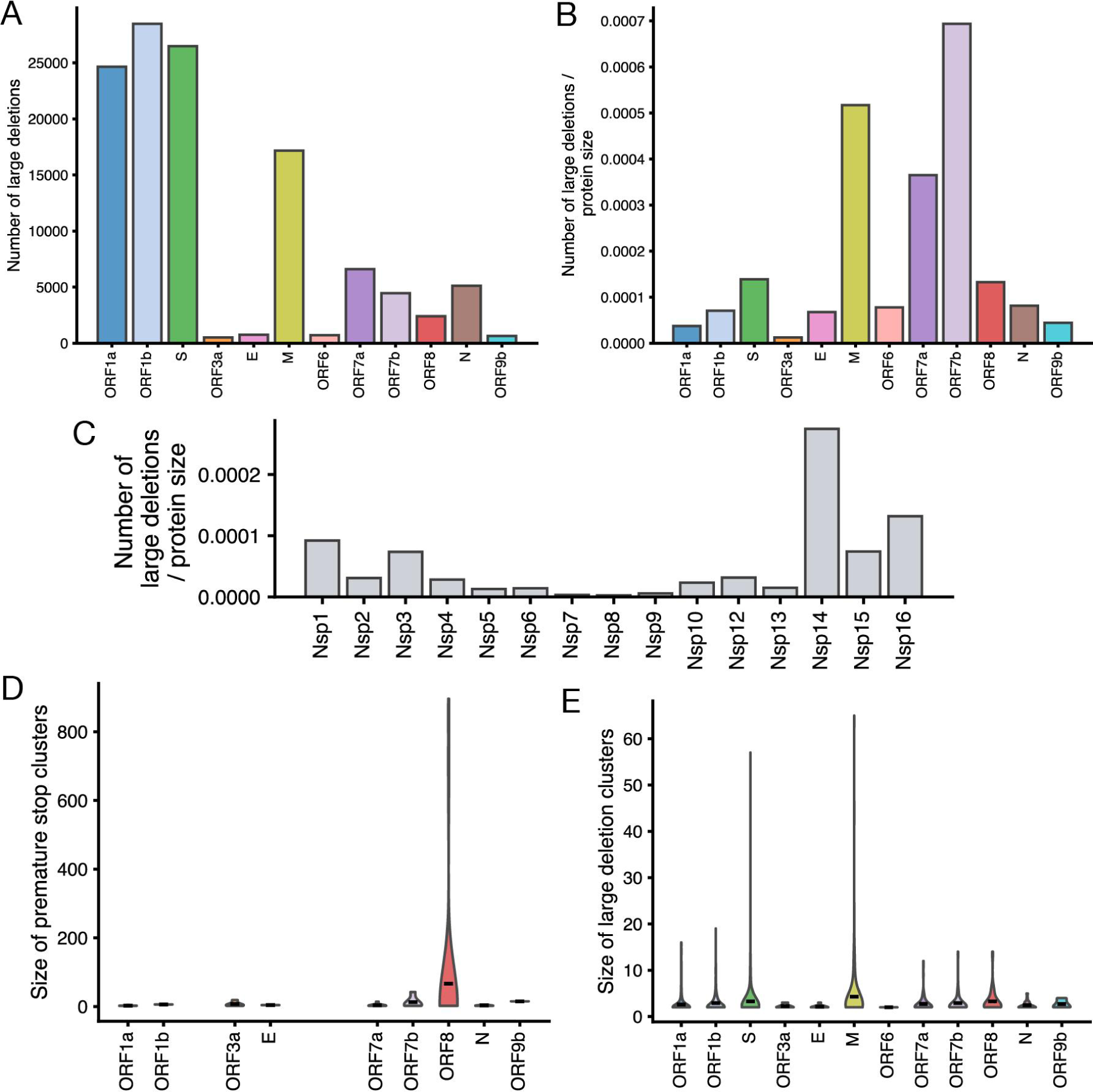
Large deletion counts by gene and reconstructed cluster sizes for gene knockouts in Washington State SARS-CoV-2 sequences. With a large deletion cutoff of 30 base pairs missing, we calculated (A) the raw number of large deletions by gene and (B) number of deletions normalized by gene length using Washington SARS-CoV-2 sequences through March 2023. (C) Using the same cutoff and dataset, we calculated the number of large deletions in ORF1a & ORF1b constituent proteins normalized by protein length. We also calculated the size of gene knockout clusters due to premature stops (D) and large deletions (E) by gene. Clusters were reconstructed from the Washington state SARS-CoV-2 phylogeny shown in Fig 1C.

**S4.**
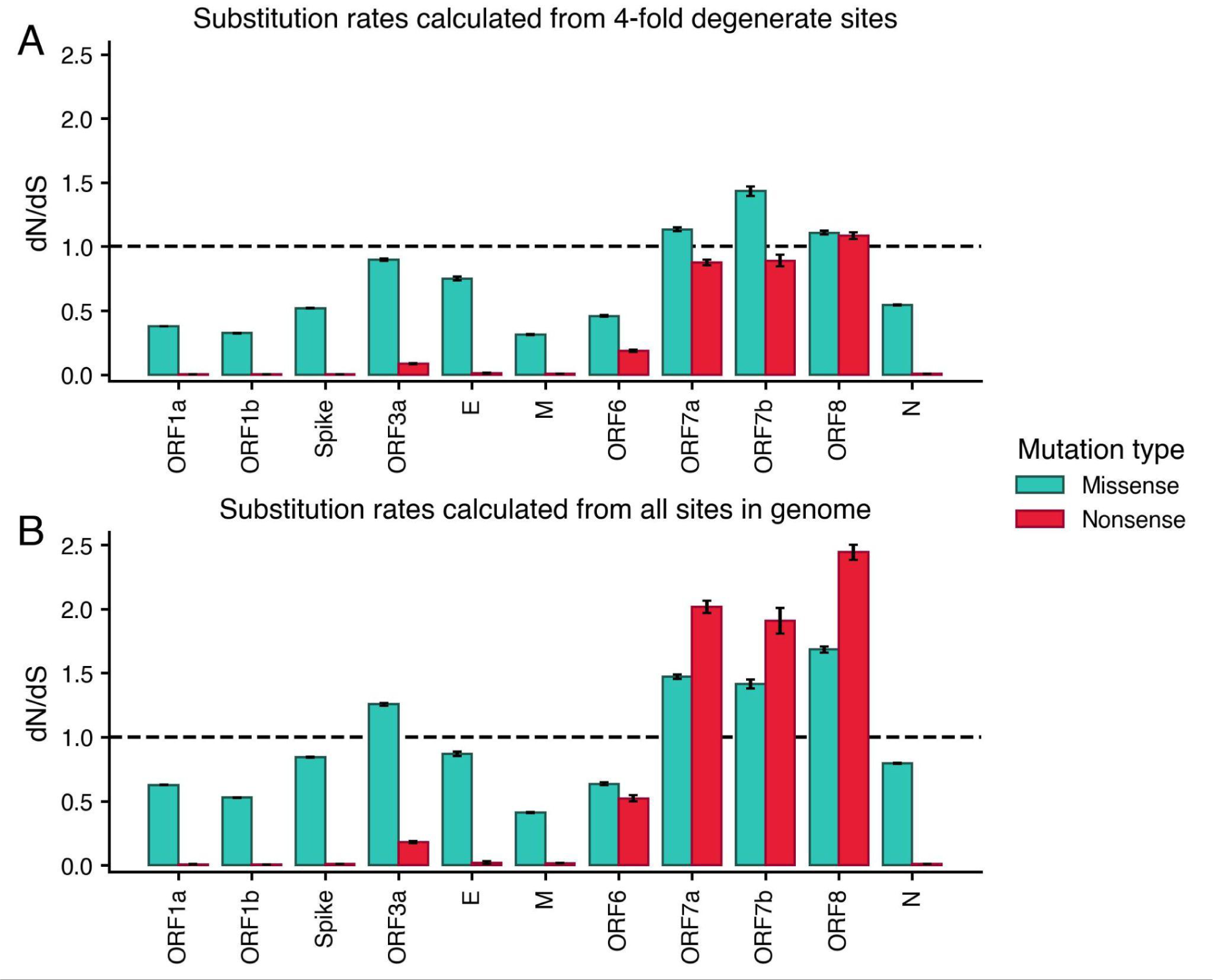
*dN/dS* values split out by mutation type for all SARS-CoV-2 genes. *dn/dS* values were calculated from the global, SARS-CoV-2 UShER phylogeny for each gene split out by missense (teal) and nonsense (red) mutations. In (A), the substitution matrix used to estimate the expected number of sites was calculated from 4-fold degenerate sites in the SARS-CoV-2 UShER phylogeny^20^. In (B), the substitution rates were inferred from all sites in the SARS-CoV-2 genome early in the pandemic^65^.

**S5.**
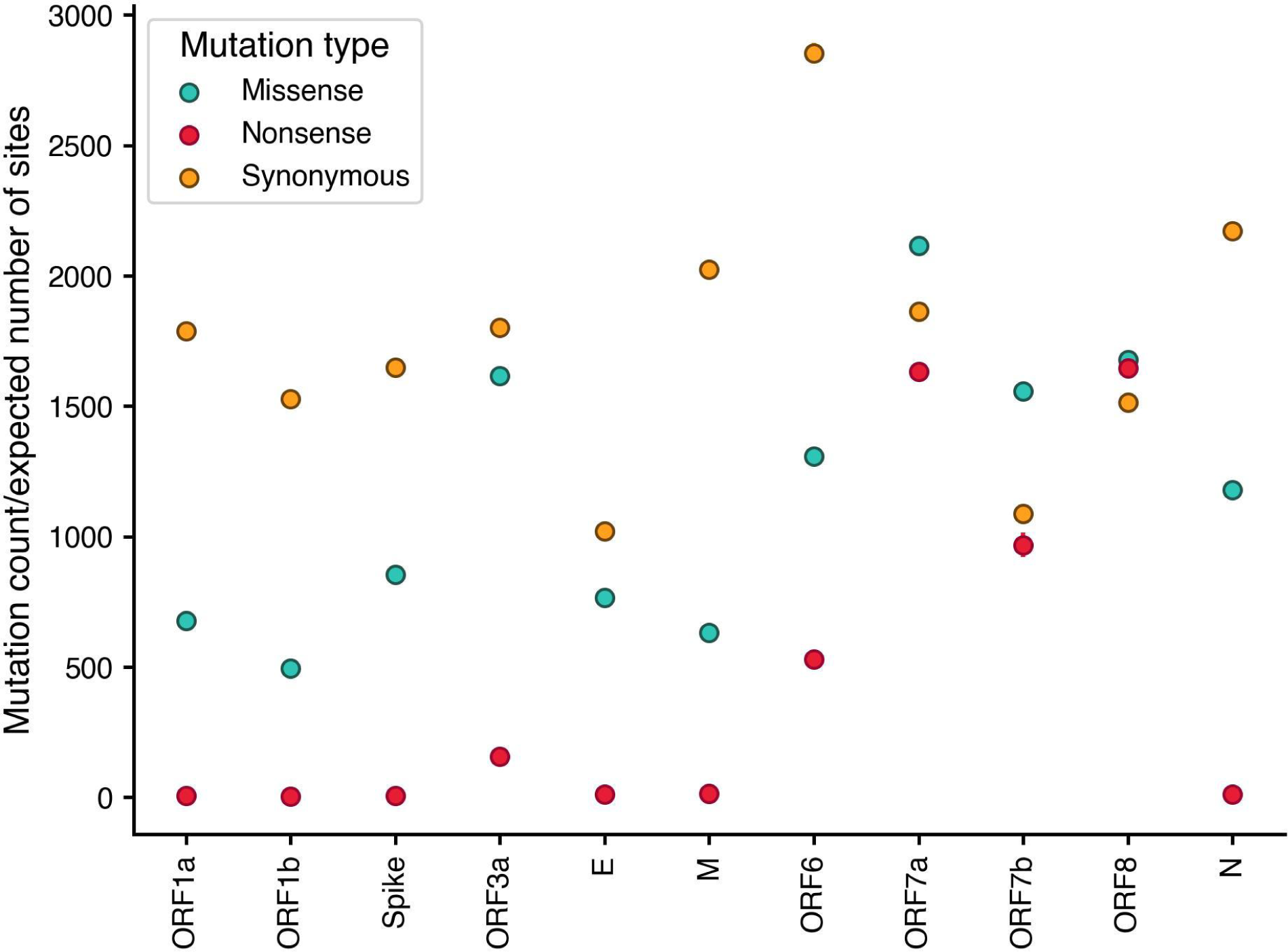
Divergence ratios for synonymous, missense and nonsense mutations for all SARS-CoV-2 genes. Divergence ratios, or the mutation count divided by the expected number of sites, were calculated from the global, SARS-CoV-2 UShER phylogeny for each gene for synonymous (yellow), missense (teal) and nonsense (red) mutations. These estimated divergence ratios correspond to *dN*, for missense and nonsense mutations, and *dS* for synonymous mutations. The expected number of sites was estimated using substitution rates inferred from the 4-fold degenerate sites.

**S6.**
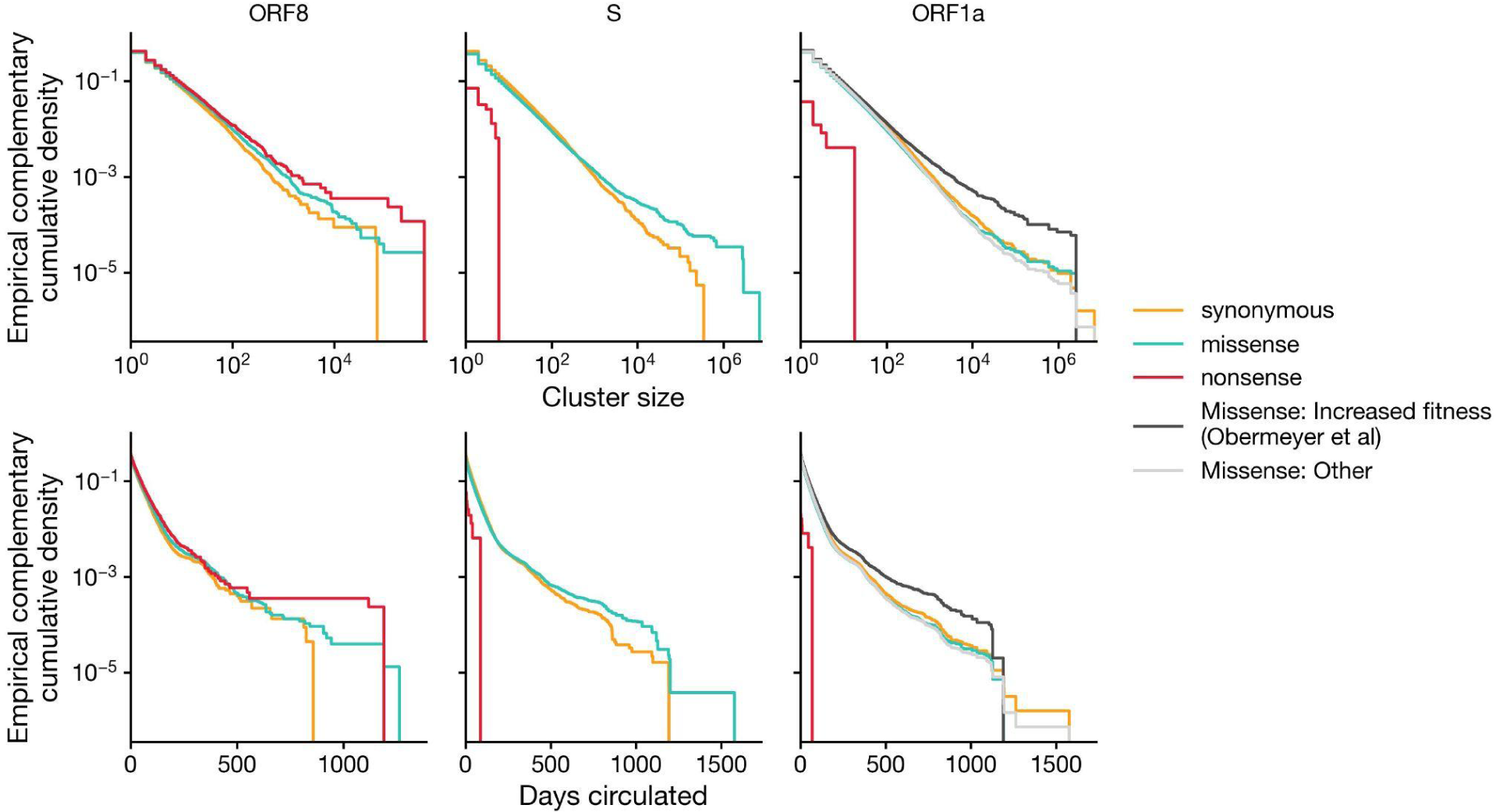
Distribution of cluster size and days circulated by mutation type for mutation clusters in ORF8, Spike and ORF1a in UShER global phylogeny. Clusters size is all descendants following a synonymous (yellow), missense (teal), or nonsense (red) mutation on the UShER SARS-CoV-2 global phylogeny. Days circulated was determined by subtracting the first date a descendant was sampled from the last date a descendant was sampled. For ORF1a, we additionally split out missense mutations into mutations associated with increased fitness (black) by a hierarchical logistic regression model developed by Obermeyer et al and all other missense mutations (silver)^19^.

**S7.**
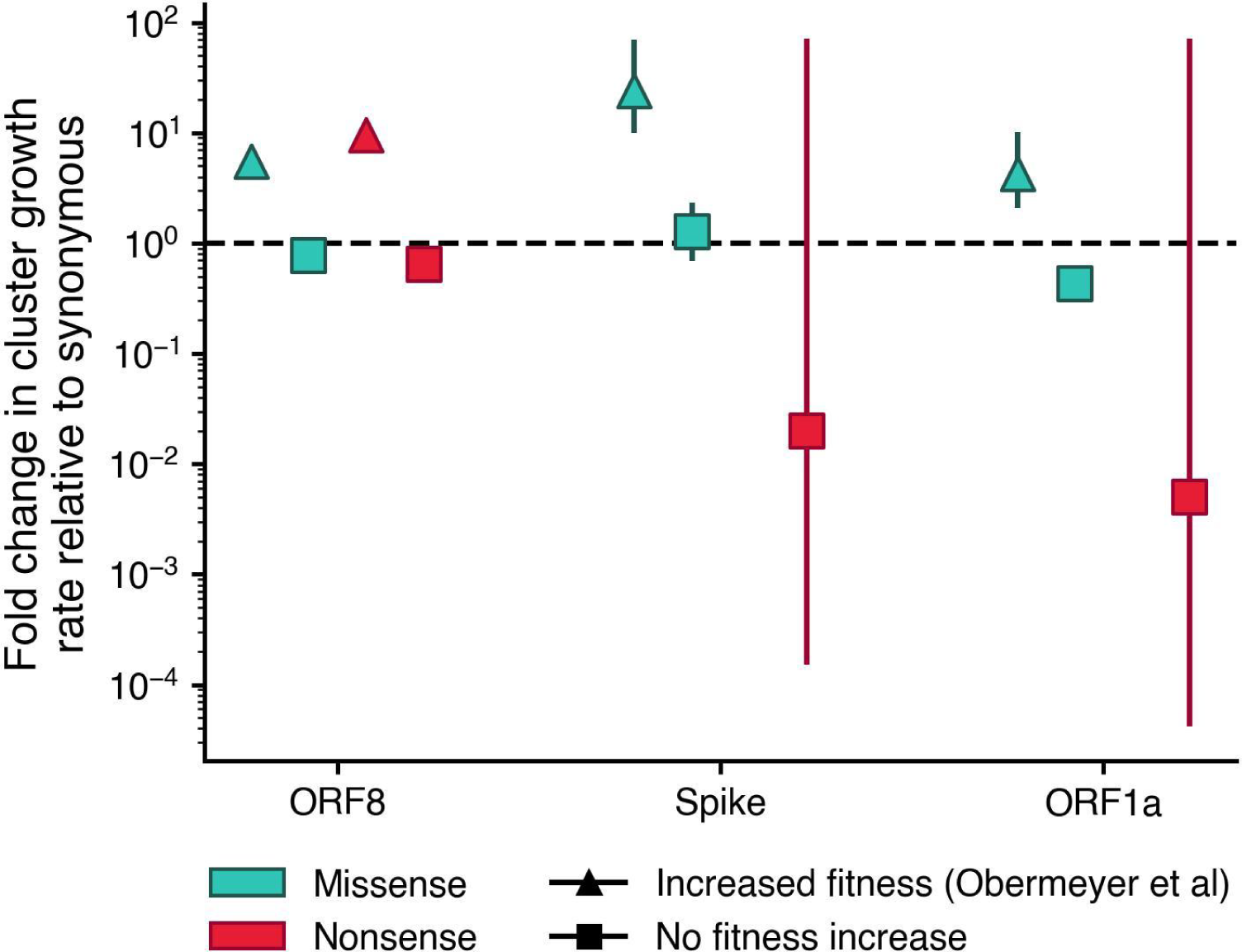
Cluster growth rates split out by mutations associated with increased fitness in Obermeyer et al. We estimated the fold change in nonsynonymous cluster growth rates relative to synonymous using negative binomial regression for ORF8, Spike, and ORF1a, splitting out mutations by those with increased fitness inferred by Obermeyer et al^19^. Nonsynonymous mutation clusters were split into four categories: missense mutations with previously inferred increased fitness (teal triangles), nonsense mutations with previously inferred increased fitness (red triangles), missense mutations without a previously inferred fitness benefit (teal squares), and nonsense mutations without a previously inferred fitness benefit (red squares). Bars indicate the 95% confidence interval.

**S8.**
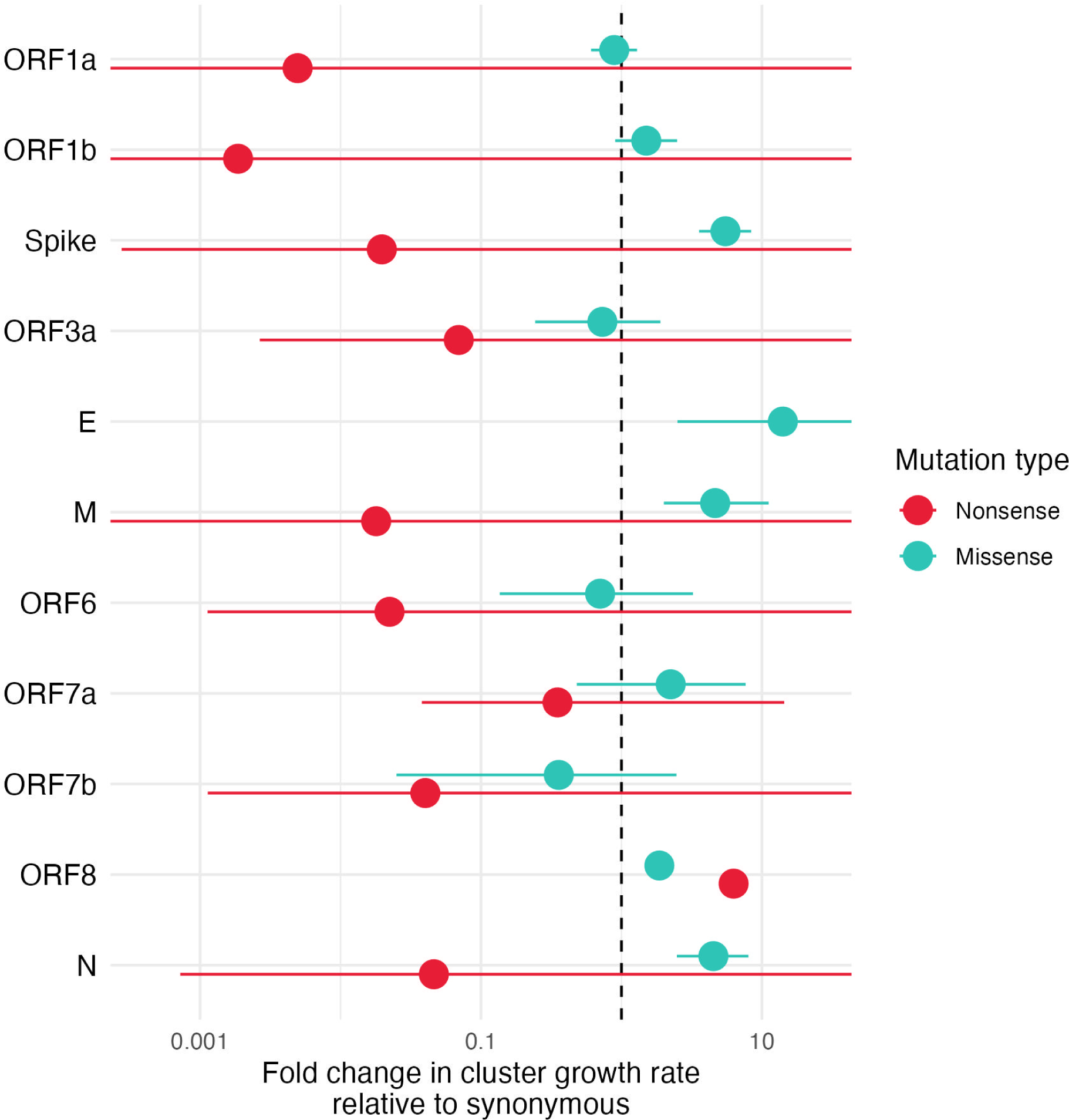
Fold change in cluster growth rate relative to synonymous for all SARS-CoV-2 genes. We estimated the fold change in cluster growth rates using a negative binomial regression for each gene in SARS-CoV-2 for clades following nonsense mutations (red) or missense mutations (teal). Bars indicate the 95% confidence interval. E did not have enough nonsense mutations to estimate a cluster growth rate.

**S9.**
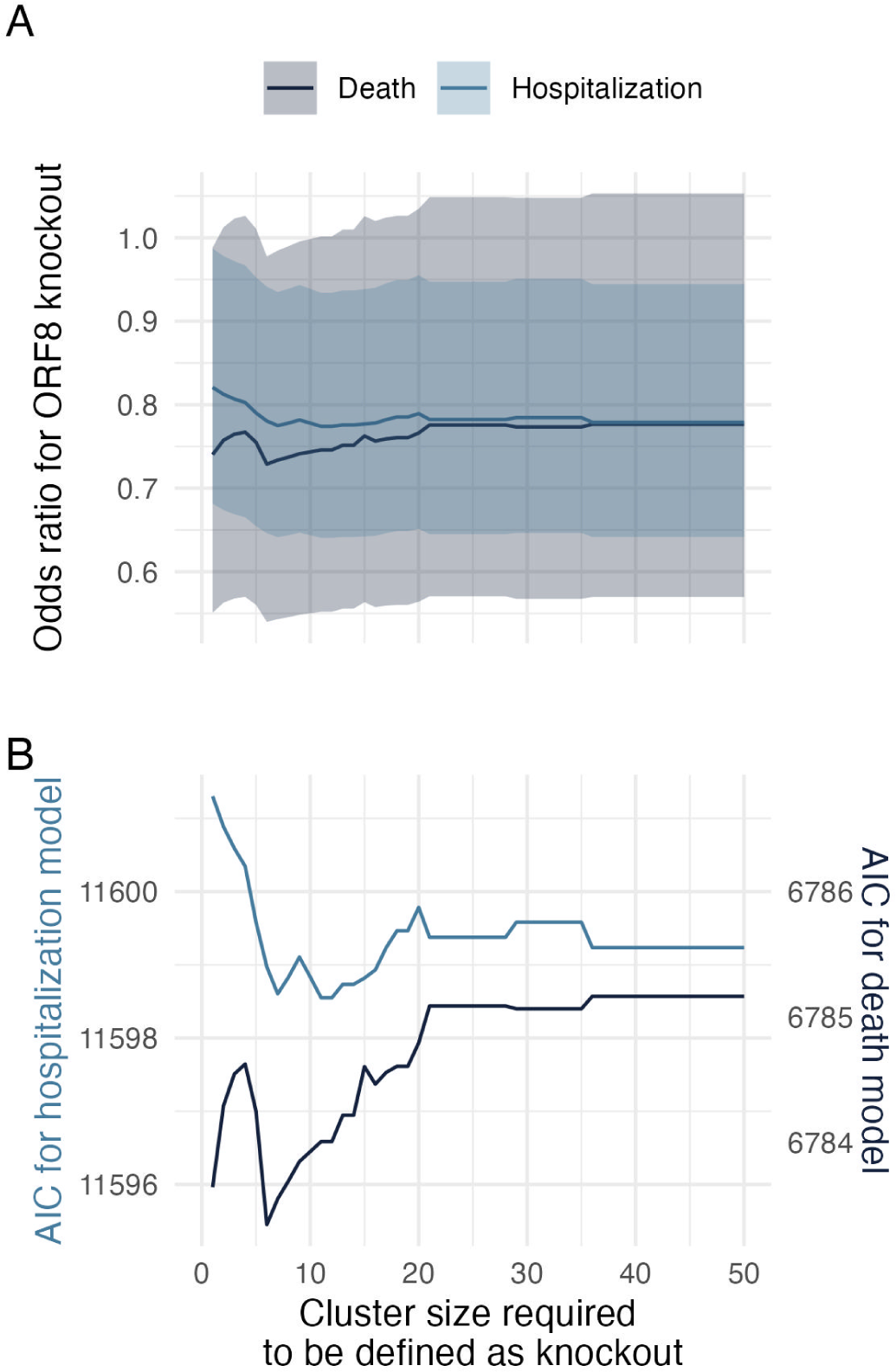
Clinical effect size of ORF8 knockout and model fit robust to cluster size. Odds ratio (A) and model Akaike Information Criterion (B) by cluster size required to define a knockout for GLMs of hospitalization (light blue) and death (dark blue). To calculate cluster size, we built three lineage-specific (Delta, Alpha, and other non-Omicron), SARS-CoV-2 maximum likelihood phylogenies enriched for ORF8 knockouts in WA and reconstructed parsimony clusters of ORF8 knockout. Breakdowns were chosen such that all ORF8 knockouts sequenced in WA could be placed in an appropriate phylogenetic context of at least 75% background sequences.

**Table S1.**
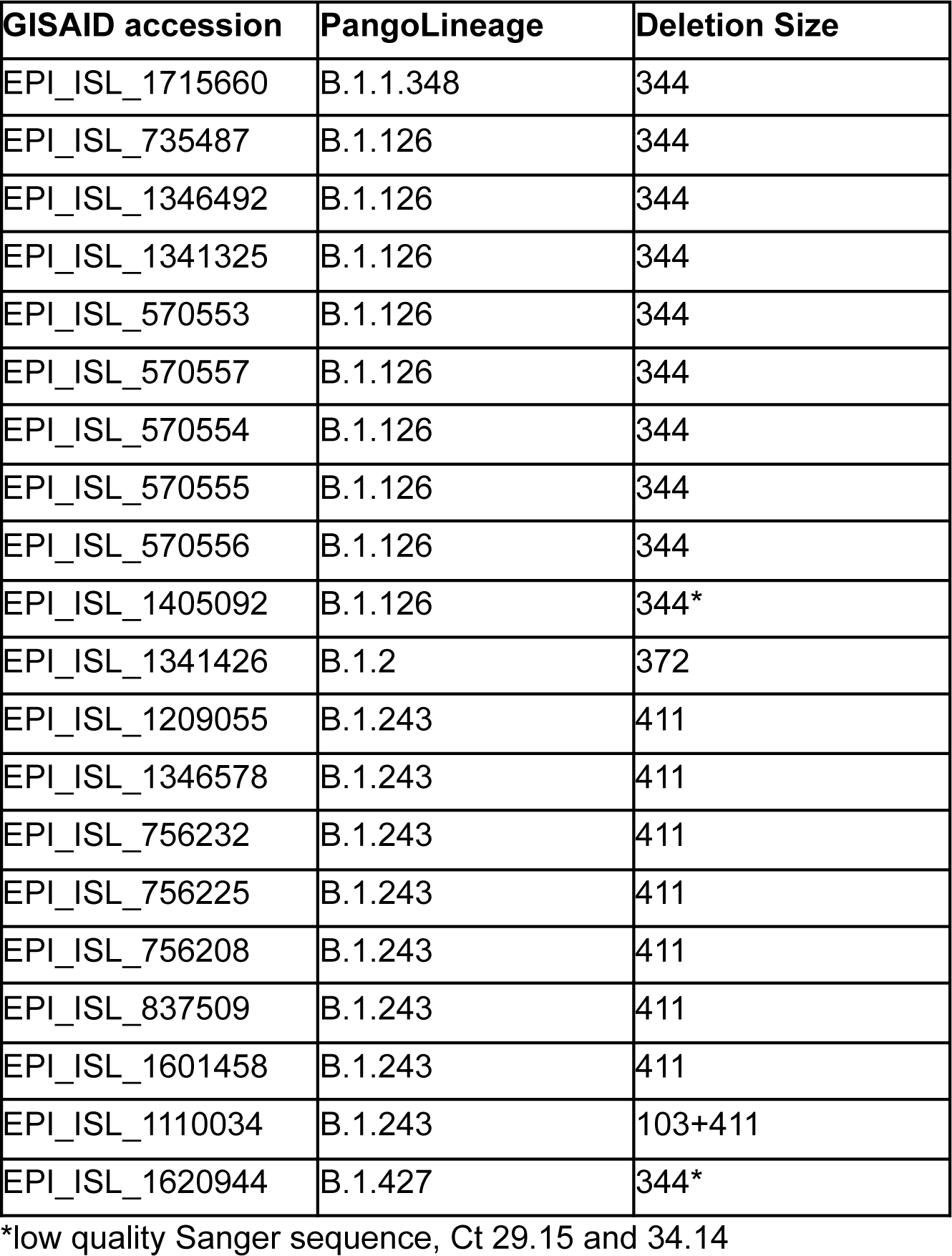
Long deletions confirmed by PCR and Sanger sequencing.

**Table S2.** Negative binomial regression of cluster size by mutation type in ORF8, excluding clusters with a G8* or Q27* mutation with log(time) observed as an offset.

| **Variable** | **Odds Ratio** | **95% CI** |
| --- | --- | --- |
| Missense mutation | 1.86 | 1.77-1.96 |
| Nonsense mutation | 2.52 | 2.30-2.76 |

**Table S3.** Clinical characteristics for sequenced SARS-CoV-2 samples in Washington Disease Reporting System stratified by ORF8 knockout.

|  | **ORF8 intact**<br>**(N=49912)** | **ORF8 knockout**<br>**(N=10089)** | **Overall**<br>**(N=60001)** |
| --- | --- | --- | --- |
| **Hospitalized?** |  |  |  |
| No | 21022 (42.1%) | 5363 (53.2%) | 26385 (44.0%) |
| Yes | 1906 (3.8%) | 383 (3.8%) | 2289 (3.8%) |
| **Missing** | 26984 (54.1%) | 4343 (43.0%) | 31327 (52.2%) |
| **Died?** |  |  |  |
| No | 49002 (98.2%) | 9960 (98.7%) | 58962 (98.3%) |
| Yes | 910 (1.8%) | 129 (1.3%) | 1039 (1.7%) |
| **Variant of concern?** |  |  |  |
| No | 10601 (21.2%) | 593 (5.9%) | 11194 (18.7%) |
| Yes | 39311 (78.8%) | 9496 (94.1%) | 48807 (81.3%) |
| **Age group** |  |  |  |
| 0-4 | 1785 (3.6%) | 431 (4.3%) | 2216 (3.7%) |
| 5-17 | 7439 (14.9%) | 1887 (18.7%) | 9326 (15.5%) |
| 18-44 | 22675 (45.4%) | 5006 (49.6%) | 27681 (46.1%) |
| 45-64 | 9677 (19.4%) | 1887 (18.7%) | 11564 (19.3%) |
| 65-79 | 3239 (6.5%) | 448 (4.4%) | 3687 (6.1%) |
| 80+ | 992 (2.0%) | 112 (1.1%) | 1104 (1.8%) |
| Missing | 4105 (8.2%) | 318 (3.2%) | 4423 (7.4%) |
| **Sex assigned at birth** |  |  |  |
| Female | 22199 (44.5%) | 4730 (46.9%) | 26929 (44.9%) |
| Male | 22757 (45.6%) | 4890 (48.5%) | 27647 (46.1%) |
| Other | 49 (0.1%) | 13 (0.1%) | 62 (0.1%) |
| Missing | 4907 (9.8%) | 456 (4.5%) | 5363 (8.9%) |
| **Vaccinated** |  |  |  |
| No | 37146 (74.4%) | 8957 (88.8%) | 46103 (76.8%) |
| Yes | 12766 (25.6%) | 1132 (11.2%) | 13898 (23.2%) |

## References

1. Plante, J.A., Liu, Y., Liu, J., Xia, H., Johnson, B.A., Lokugamage, K.G., Zhang, X., Muruato, A.E., Zou, J., Fontes-Garfias, C.R., et al. (2021). Spike mutation D614G alters SARS-CoV-2 fitness. Nature 592, 116–121.

2. Korber, B., Fischer, W.M., Gnanakaran, S., Yoon, H., Theiler, J., Abfalterer, W., Hengartner, N., Giorgi, E.E., Bhattacharya, T., Foley, B., et al. (2020). Tracking Changes in SARS-CoV-2 Spike: Evidence that D614G Increases Infectivity of the COVID-19 Virus. Cell 182, 812–827.e19.

3. Andrew, R. (2020). Preliminary genomic characterisation of an emergent SARS-CoV-2 lineage in the UK defined by a novel set of spike mutations. virological.

4. Tegally, H., Wilkinson, E., Giovanetti, M., Iranzadeh, A., Fonseca, V., Giandhari, J., Doolabh, D., Pillay, S., San, E.J., Msomi, N., et al. (2021). Detection of a SARS-CoV-2 variant of concern in South Africa. Nature 592, 438–443.

5. Faria, N.R., Mellan, T.A., Whittaker, C., Claro, I.M., Candido, D. da S., Mishra, S., Crispim, M.A.E., Sales, F.C.S., Hawryluk, I., McCrone, J.T., et al. (2021). Genomics and epidemiology of the P.1 SARS-CoV-2 lineage in Manaus, Brazil. Science 372, 815–821.

6. Tao, K., Tzou, P.L., Nouhin, J., Gupta, R.K., de Oliveira, T., Kosakovsky Pond, S.L., Fera, D., and Shafer, R.W. (2021). The biological and clinical significance of emerging SARS-CoV-2 variants. Nat. Rev. Genet. 22, 757–773.

7. Liu, Y., Liu, J., Johnson, B.A., Xia, H., Ku, Z., Schindewolf, C., Widen, S.G., An, Z., Weaver, S.C., Menachery, V.D., et al. (2022). Delta spike P681R mutation enhances SARS-CoV-2 fitness over Alpha variant. Cell Rep. 39, 110829.

8. Mlcochova, P., Kemp, S.A., Dhar, M.S., Papa, G., Meng, B., Ferreira, I.A.T.M., Datir, R., Collier, D.A., Albecka, A., Singh, S., et al. (2021). SARS-CoV-2 B.1.617.2 Delta variant replication and immune evasion. Nature 599, 114–119.

9. Tegally, H., Moir, M., Everatt, J., Giovanetti, M., Scheepers, C., Wilkinson, E., Subramoney, K., Makatini, Z., Moyo, S., Amoako, D.G., et al. (2022). Emergence of SARS-CoV-2 Omicron lineages BA.4 and BA.5 in South Africa. Nat. Med. 28, 1785–1790.

10. Ito, K., Piantham, C., and Nishiura, H. (2022). Estimating relative generation times and reproduction numbers of Omicron BA.1 and BA.2 with respect to Delta variant in Denmark. Math. Biosci. Eng. 19, 9005–9017.

11. Tamura, T., Ito, J., Uriu, K., Zahradnik, J., Kida, I., Anraku, Y., Nasser, H., Shofa, M., Oda, Y., Lytras, S., et al. (2023). Virological characteristics of the SARS-CoV-2 XBB variant derived from recombination of two Omicron subvariants. Nat. Commun. 14, 2800.

12. Ozono, S., Zhang, Y., Ode, H., Sano, K., Tan, T.S., Imai, K., Miyoshi, K., Kishigami, S., Ueno, T., Iwatani, Y., et al. (2021). SARS-CoV-2 D614G spike mutation increases entry efficiency with enhanced ACE2-binding affinity. Nat. Commun. 12, 848.

13. Zhou, B., Thao, T.T.N., Hoffmann, D., Taddeo, A., Ebert, N., Labroussaa, F., Pohlmann, A., King, J., Steiner, S., Kelly, J.N., et al. (2021). SARS-CoV-2 spike D614G change enhances replication and transmission. Nature 592, 122–127.

14. Starr, T.N., Greaney, A.J., Hannon, W.W., Loes, A.N., Hauser, K., Dillen, J.R., Ferri, E., Farrell, A.G., Dadonaite, B., McCallum, M., et al. (2022). Shifting mutational constraints in the SARS-CoV-2 receptor-binding domain during viral evolution. Science 377, 420–424.

15. Saito, A., Irie, T., Suzuki, R., Maemura, T., Nasser, H., Uriu, K., Kosugi, Y., Shirakawa, K., Sadamasu, K., Kimura, I., et al. (2022). Enhanced fusogenicity and pathogenicity of SARS-CoV-2 Delta P681R mutation. Nature 602, 300–306.

16. Yue, C., Song, W., Wang, L., Jian, F., Chen, X., Gao, F., Shen, Z., Wang, Y., Wang, X., and Cao, Y. (2023). ACE2 binding and antibody evasion in enhanced transmissibility of XBB.1.5. Lancet Infect. Dis. 23, 278–280.

17. Jackson, C.B., Farzan, M., Chen, B., and Choe, H. (2022). Mechanisms of SARS-CoV-2 entry into cells. Nat. Rev. Mol. Cell Biol. 23, 3–20.

18. Kistler, K.E., Huddleston, J., and Bedford, T. (2022). Rapid and parallel adaptive mutations in spike S1 drive clade success in SARS-CoV-2. Cell Host Microbe 30, 545–555.e4.

19. Obermeyer, F., Jankowiak, M., Barkas, N., Schaffner, S.F., Pyle, J.D., Yurkovetskiy, L., Bosso, M., Park, D.J., Babadi, M., MacInnis, B.L., et al. (2022). Analysis of 6.4 million SARS-CoV-2 genomes identifies mutations associated with fitness. Science 376, 1327–1332.

20. Bloom, J.D., and Neher, R.A. (2023). Fitness effects of mutations to SARS-CoV-2 proteins. bioRxiv. 10.1101/2023.01.30.526314.

21. Su, Y.C.F., Anderson, D.E., Young, B.E., Linster, M., Zhu, F., Jayakumar, J., Zhuang, Y., Kalimuddin, S., Low, J.G.H., Tan, C.W., et al. (2020). Discovery and Genomic Characterization of a 382-Nucleotide Deletion in ORF7b and ORF8 during the Early Evolution of SARS-CoV-2. MBio 11. 10.1128/mBio.01610-20.

22. Young, B.E., Fong, S.-W., Chan, Y.-H., Mak, T.-M., Ang, L.W., Anderson, D.E., Lee, C.Y.-P., Amrun, S.N., Lee, B., Goh, Y.S., et al. (2020). Effects of a major deletion in the SARS-CoV-2 genome on the severity of infection and the inflammatory response: an observational cohort study. Lancet 396, 603–611.

23. Gong, Y.-N., Tsao, K.-C., Hsiao, M.-J., Huang, C.-G., Huang, P.-N., Huang, P.-W., Lee, K.-M., Liu, Y.-C., Yang, S.-L., Kuo, R.-L., et al. (2020). SARS-CoV-2 genomic surveillance in Taiwan revealed novel ORF8-deletion mutant and clade possibly associated with infections in Middle East. Emerg. Microbes Infect. 9, 1457–1466.

24. Saha, O., Hossain, M.S., and Rahaman, M.M. (2020). Genomic exploration light on multiple origin with potential parsimony-informative sites of the severe acute respiratory syndrome coronavirus 2 in Bangladesh. Gene Rep 21, 100951.

25. Mazur-Panasiuk, N., Rabalski, L., Gromowski, T., Nowicki, G., Kowalski, M., Wydmanski, W., Szulc, P., Kosinski, M., Gackowska, K., Drweska-Matelska, N., et al. (2021). Expansion of a SARS-CoV-2 Delta variant with an 872 nt deletion encompassing ORF7a, ORF7b and ORF8, Poland, July to August 2021. Euro Surveill. 26. 10.2807/1560-7917.ES.2021.26.39.2100902.

26. Ko, K., Nagashima, S., E, B., Ouoba, S., Akita, T., Sugiyama, A., Ohisa, M., Sakaguchi, T., Tahara, H., Ohge, H., et al. (2021). Molecular characterization and the mutation pattern of SARS-CoV-2 during first and second wave outbreaks in Hiroshima, Japan. PLoS One 16, e0246383.

27. Pereira, F. (2021). SARS-CoV-2 variants lacking ORF8 occurred in farmed mink and pangolin. Gene 784, 145596.

28. DeRonde, S., Deuling, H., Parker, J., and Chen, J. (2022). Identification of a novel SARS-CoV-2 variant with a truncated protein in ORF8 gene by next generation sequencing. Sci. Rep. 12, 4631.

29. auspice https://nextstrain.org/ncov/gisaid/global/all-time.

30. Chinese SARS Molecular Epidemiology Consortium (2004). Molecular evolution of the SARS coronavirus during the course of the SARS epidemic in China. Science 303, 1666–1669.

31. Arduini, A., Laprise, F., and Liang, C. (2023). SARS-CoV-2 ORF8: A Rapidly Evolving Immune and Viral Modulator in COVID-19. Viruses 15. 10.3390/v15040871.

32. Hachim, A., Kavian, N., Cohen, C.A., Chin, A.W.H., Chu, D.K.W., Mok, C.K.P., Tsang, O.T.Y., Yeung, Y.C., Perera, R.A.P.M., Poon, L.L.M., et al. (2020). ORF8 and ORF3b antibodies are accurate serological markers of early and late SARS-CoV-2 infection. Nat. Immunol. 21, 1293–1301.

33. Matsuoka, K., Imahashi, N., Ohno, M., Ode, H., Nakata, Y., Kubota, M., Sugimoto, A., Imahashi, M., Yokomaku, Y., and Iwatani, Y. (2022). SARS-CoV-2 accessory protein ORF8 is secreted extracellularly as a glycoprotein homodimer. J. Biol. Chem. 298, 101724.

34. Zhang, Y., Chen, Y., Li, Y., Huang, F., Luo, B., Yuan, Y., Xia, B., Ma, X., Yang, T., Yu, F., et al. (2021). The ORF8 protein of SARS-CoV-2 mediates immune evasion through down-regulating MHC-Ι. Proc. Natl. Acad. Sci. U. S. A. 118. 10.1073/pnas.2024202118.

35. Moriyama, M., Lucas, C., Monteiro, V.S., Yale SARS-CoV-2 Genomic Surveillance Initiative, and Iwasaki, A. (2022). SARS-CoV-2 Omicron subvariants evolved to promote further escape from MHC-I recognition. bioRxiv. 10.1101/2022.05.04.490614.

36. Beaudoin-Bussières, G., Arduini, A., Bourassa, C., Medjahed, H., Gendron-Lepage, G., Richard, J., Pan, Q., Wang, Z., Liang, C., and Finzi, A. (2022). SARS-CoV-2 Accessory Protein ORF8 Decreases Antibody-Dependent Cellular Cytotoxicity. Viruses 14. 10.3390/v14061237.

37. Li, J.-Y., Liao, C.-H., Wang, Q., Tan, Y.-J., Luo, R., Qiu, Y., and Ge, X.-Y. (2020). The ORF6, ORF8 and nucleocapsid proteins of SARS-CoV-2 inhibit type I interferon signaling pathway. Virus Res. 286, 198074.

38. Rashid, F., Dzakah, E.E., Wang, H., and Tang, S. (2021). The ORF8 protein of SARS-CoV-2 induced endoplasmic reticulum stress and mediated immune evasion by antagonizing production of interferon beta. Virus Res. 296, 198350.

39. Chen, J., Lu, Z., Yang, X., Zhou, Y., Gao, J., Zhang, S., Huang, S., Cai, J., Yu, J., Zhao, W., et al. (2022). Severe Acute Respiratory Syndrome Coronavirus 2 ORF8 Protein Inhibits Type I Interferon Production by Targeting HSP90B1 Signaling. Front. Cell. Infect. Microbiol. 12, 899546.

40. Rashid, F., Suleman, M., Shah, A., Dzakah, E.E., Wang, H., Chen, S., and Tang, S. (2021). Mutations in SARS-CoV-2 ORF8 Altered the Bonding Network With Interferon Regulatory Factor 3 to Evade Host Immune System. Front. Microbiol. 12, 703145.

41. Geng, H., Subramanian, S., Wu, L., Bu, H.-F., Wang, X., Du, C., De Plaen, I.G., and Tan, X.-D. (2021). SARS-CoV-2 ORF8 Forms Intracellular Aggregates and Inhibits IFNγ-Induced Antiviral Gene Expression in Human Lung Epithelial Cells. Front. Immunol. 12, 679482.

42. Kee, J., Thudium, S., Renner, D.M., Glastad, K., Palozola, K., Zhang, Z., Li, Y., Lan, Y., Cesare, J., Poleshko, A., et al. (2022). SARS-CoV-2 disrupts host epigenetic regulation via histone mimicry. Nature 610, 381–388.

43. Lin, X., Fu, B., Yin, S., Li, Z., Liu, H., Zhang, H., Xing, N., Wang, Y., Xue, W., Xiong, Y., et al. (2021). ORF8 contributes to cytokine storm during SARS-CoV-2 infection by activating IL-17 pathway. iScience 24, 102293.

44. Wu, X., Xia, T., Shin, W.-J., Yu, K.-M., Jung, W., Herrmann, A., Foo, S.-S., Chen, W., Zhang, P., Lee, J.-S., et al. (2022). Viral Mimicry of Interleukin-17A by SARS-CoV-2 ORF8. MBio 13, e0040222.

45. Lin, X., Fu, B., Xiong, Y., Xing, N., Xue, W., Guo, D., Zaky, M., Pavani, K., Kunec, D., Trimpert, J., et al. (2023). Unconventional secretion of unglycosylated ORF8 is critical for the cytokine storm during SARS-CoV-2 infection. PLoS Pathog. 19, e1011128.

46. Bedford, T., Greninger, A.L., Roychoudhury, P., Starita, L.M., Famulare, M., Huang, M.-L., Nalla, A., Pepper, G., Reinhardt, A., Xie, H., et al. (2020). Cryptic transmission of SARS-CoV-2 in Washington state. Science 370, 571–575.

47. Paredes, M.I., Lunn, S.M., Famulare, M., Frisbie, L.A., Painter, I., Burstein, R., Roychoudhury, P., Xie, H., Mohamed Bakhash, S.A., Perez, R., et al. (2022). Associations Between Severe Acute Respiratory Syndrome Coronavirus 2 (SARS-CoV-2) Variants and Risk of Coronavirus Disease 2019 (COVID-19) Hospitalization Among Confirmed Cases in Washington State: A Retrospective Cohort Study. Clin. Infect. Dis. 75, e536–e544.

48. Oltean, H.N., Allen, K.J., Frisbie, L., Lunn, S.M., Torres, L.M., Manahan, L., Painter, I., Russell, D., Singh, A., Peterson, J.M., et al. (2023). Sentinel Surveillance System Implementation and Evaluation for SARS-CoV-2 Genomic Data, Washington, USA, 2020-2021. Emerg. Infect. Dis. 29, 242–251.

49. Washington State Department of Health (2023). SARS-CoV-2 Sequencing and Variants Report.

50. Turakhia, Y., Thornlow, B., Hinrichs, A.S., De Maio, N., Gozashti, L., Lanfear, R., Haussler, D., and Corbett-Detig, R. (2021). Ultrafast Sample placement on Existing tRees (UShER) enables real-time phylogenetics for the SARS-CoV-2 pandemic. Nat. Genet. 53, 809–816.

51. McBroome, J., Thornlow, B., Hinrichs, A.S., Kramer, A., De Maio, N., Goldman, N., Haussler, D., Corbett-Detig, R., and Turakhia, Y. (2021). A Daily-Updated Database and Tools for Comprehensive SARS-CoV-2 Mutation-Annotated Trees. Mol. Biol. Evol. 38, 5819–5824.

52. Kryazhimskiy, S., and Plotkin, J.B. (2008). The population genetics of dN/dS. PLoS Genet. 4, e1000304.

53. Mugal, C.F., Wolf, J.B.W., and Kaj, I. (2014). Why time matters: codon evolution and the temporal dynamics of dN/dS. Mol. Biol. Evol. 31, 212–231.

54. Addetia, A., Xie, H., Roychoudhury, P., Shrestha, L., Loprieno, M., Huang, M.-L., Jerome, K.R., and Greninger, A.L. (2020). Identification of multiple large deletions in ORF7a resulting in in-frame gene fusions in clinical SARS-CoV-2 isolates. J. Clin. Virol. 129, 104523.

55. Kim, I.-J., Lee, Y.-H., Khalid, M.M., Chen, I.P., Zhang, Y., Ott, M., and Verdin, E. (2023). SARS-CoV-2 protein ORF8 limits expression levels of Spike antigen and facilitates immune evasion of infected host cells. J. Biol. Chem. 299, 104955.

56. Shu, Y., and McCauley, J. (2017). GISAID: Global initiative on sharing all influenza data - from vision to reality. Euro Surveill. 22. 10.2807/1560-7917.ES.2017.22.13.30494.

57. Müller, N.F., Wagner, C., Frazar, C.D., Roychoudhury, P., Lee, J., Moncla, L.H., Pelle, B., Richardson, M., Ryke, E., Xie, H., et al. (2021). Viral genomes reveal patterns of the SARS-CoV-2 outbreak in Washington State. Sci. Transl. Med. 13. 10.1126/scitranslmed.abf0202.

58. Ulhuq, F.R., Barge, M., Falconer, K., Wild, J., Fernandes, G., Gallagher, A., McGinley, S., Sugadol, A., Tariq, M., Maloney, D., et al. (2023). Analysis of the ARTIC V4 and V4.1 SARS-CoV-2 primers and their impact on the detection of Omicron BA.1 and BA.2 lineage-defining mutations. Microb Genom 9. 10.1099/mgen.0.000991.

59. Hadfield, J., Megill, C., Bell, S.M., Huddleston, J., Potter, B., Callender, C., Sagulenko, P., Bedford, T., and Neher, R.A. (2018). Nextstrain: real-time tracking of pathogen evolution. Bioinformatics 34, 4121–4123.

60. Aksamentov, I., Roemer, C., Hodcroft, E., and Neher, R. (2021). Nextclade: clade assignment, mutation calling and quality control for viral genomes. J. Open Source Softw. 6, 3773.

61. Minh, B.Q., Schmidt, H.A., Chernomor, O., Schrempf, D., Woodhams, M.D., von Haeseler, A., and Lanfear, R. (2020). Corrigendum to: IQ-TREE 2: New Models and Efficient Methods for Phylogenetic Inference in the Genomic Era. Mol. Biol. Evol. 37, 2461.

62. Sagulenko, P., Puller, V., and Neher, R.A. (2018). TreeTime: Maximum-likelihood phylodynamic analysis. Virus Evol 4, vex042.

63. Camin, J.H., and Sokal, R.R. (1965). A Method for Deducing Branching Sequences in Phylogeny. Evolution 19, 311–326.

64. Fitch, W.M. (1971). Toward Defining the Course of Evolution: Minimum Change for a Specific Tree Topology. Syst. Zool. 20, 406–416.

65. Uddin, M.B., Sajib, E.H., Hoque, S.F., Bappy, M.N.I., Elahi, F., Ghosh, A., Muhit, S., Hassan, M.M., Hasan, M., Chelliah, R., et al. (2022). Genomic diversity and molecular dynamics interaction on mutational variances among RB domains of SARS-CoV-2 interplay drug inactivation. Infect. Genet. Evol. 97, 105128.

66. Updated working definitions and primary actions for SARSCoV2 variants https://www.who.int/publications/m/item/historical-working-definitions-and-primary-actions-for-sars-cov-2-variants.

